# Bi-directional Mendelian randomization and multi-phenotype GWAS show causality and shared pathophysiology between depression and type 2 diabetes

**DOI:** 10.1101/2022.12.06.22283143

**Authors:** Jared G Maina, Zhanna Balkhiyarova, Arie Nouwen, Igor Pupko, Anna Ulrich, Mathilde Boissel, Amélie Bonnefond, Philippe Froguel, Amna Khamis, Inga Prokopenko, Marika Kaakinen

**Author notes:** Corresponding authors: Dr Marika Kaakinen, Section of Statistical Multi-Omics, Department of Clinical and Experimental Medicine, University of Surrey, Leggett Building, Daphne Jackson Road, Manor Campus, Guildford, Surrey, UK, GU2 7WG, Prof. Inga Prokopenko, Section of Statistical Multi-Omics, Department of Clinical and Experimental Medicine, University of Surrey, Leggett Building, Daphne Jackson Road, Manor Campus, Guildford, Surrey, UK, GU2 7WG, Dr Amna Khamis, European Genomic Institute for Diabetes - UMR 1283/8199, Pôle Recherche - 1er étage Aile Ouest, 1, Place de Verdun, 59045 LILLE CEDEX. These authors contributed equally to this research (Jared G Maina, Zhanna Balkhiyarova, Inga Prokopenko, Marika Kaakinen).

## Abstract

**OBJECTIVE:** Depression is a common co-morbidity of type 2 diabetes. However, the causality and underlying mechanisms remain unclear.

**RESEARCH DESIGN AND METHODS:** We applied bi-directional Mendelian randomization (MR) to assess causality between type 2 diabetes and self-reported depression. Using the UK biobank, we performed 1) GWAS, separately, and 2) multi-phenotype GWAS (MP-GWAS) of type 2 diabetes (cases=19,344, controls=463,641) and depression, using two depression definitions–clinically diagnosed major depressive disorder (MDD, cases=5,262, controls=86,275) and self-reported depressive symptoms (PHQ-9, n=153,079). The FinnGen study was used for replication for MDD (n=23,424) and type 2 diabetes (n=32,469). Based on the results, we analyzed expression quantitative trait loci (eQTL) data from public databases to identify target genes in relevant tissues.

**RESULTS:** MR demonstrated a significant causal effect of depression on type 2 diabetes (OR=1.18[1.06-1.32], p=0.0024), but not in the reverse direction. GWAS of type 2 diabetes and depressive symptoms did not identify any shared loci between them, whereas MP-GWAS identified seven shared loci mapped to *TCF7L2, CDKAL1, IGF2BP2, SPRY2, CCND2-AS1, IRS1, CDKN2B-AS1*. MDD did not yield genome-wide significant loci in either GWAS or MP-GWAS. We found that most MP-GWAS *loci* had an eQTL, including SNPs implicating the cell cycle gene *CCND2* in pancreatic islets and brain, and key insulin signaling gene *IRS1* in adipose tissue, suggesting a multi-tissue and pleiotropic underlying mechanism.

**CONCLUSION:** Our study reveals the complexity in the depression-diabetes relationship and our results have important implications for a more efficient prevention of type 2 diabetes from early adulthood when depressive symptoms usually occur.

## INTRODUCTION

Type 2 diabetes is a disease characterized by chronic hyperglycemia and depression is a common co-morbidity, potentially due to common shared risk factors, such as lifestyle, early growth environment, psychotropic drugs, and neuro-endocrine dysfunction^1^. Whether the impact of type 2 diabetes on depression is stronger than the reverse remains to be defined. It has indeed been shown that depression, even at sub-clinical levels, increases the risk of incident type 2 diabetes by 25-60%^2,3^, whereas others have shown that type 2 diabetes increases the risk of depression by 40-60%^4^.

The causality of the associations from observational studies remains unclear due to unmeasured confounding and potential reverse causation. However, this could be circumvented in part through Mendelian randomization, an approach that assesses potential causality between phenotypes using genetic variants as instruments, since genes are allocated randomly at birth and are free of confounding^5^. To date, only one MR study from China has reported a possible causal link from type 2 diabetes to depression^6^, yet the reverse direction of causal link was not examined.

Recent large-scale genome-wide association studies (GWAS) for type 2 diabetes and depression have reported 403 and 102 associated genomic *loci* for these diseases, respectively^7,8^. Moreover, analyses based on the GWAS results support a positive genetic correlation (r_G_) between them^7,8^, suggesting shared genetic background. However, the majority of GWAS investigate each disease independently, without considering the genetic correlation between related phenotypes and their heritabilities.

Therefore, in this study, we addressed the causal relationship between depression and type 2 diabetes by conducting an MR study using summary statistics from recent GWAS of depression^8^ and type 2 diabetes^7^. Additionally, we used the UK biobank (UKBB) to perform a multi-phenotype GWAS (MP-GWAS) for type 2 diabetes and depression to identify shared genetic *loci* between the two diseases. For depression, we compared two assessment approaches - clinically diagnosed major depressive disorder (MDD) and depressive symptoms based on self-report.

## RESEARCH DESIGN AND METHODS

### Mendelian randomization

#### Summary statistics used

To test for causality between type 2 diabetes and depression, we performed a two-sample bidirectional MR, first using depression as a risk factor and type 2 diabetes as an outcome, then testing type 2 diabetes as a risk factor and depression as an outcome from two non-overlapping datasets (**Supplementary Figure 1**). The single nucleotide polymorphisms (SNPs) used as genetic instruments for type 2 diabetes and self-reported depression were from recent large-scale European GWAS meta-analyses of the two diseases^7,8^.

#### Two-sample bidirectional MR

All MR analyses were conducted using the R software package *TwoSampleMR v0*.*5*.*4*^9^. In the type 2 diabetes GWAS summary statistics used, 95 single nucleotide polymorphisms (SNPs, inclusive of 6 proxy variants with a minimum r^2^≥0.8) out of the 102 independent (r^2^<0.01) depression SNPs were available. We excluded 7 palindromic SNPs (A/T or C/G) with intermediate allele frequencies (minor allele frequency, MAF>45%) to ensure the effects of the SNPs for the two phenotypes were aligned to the same forward strand allele. The genetic instruments for type 2 diabetes included 403 genetic SNPs associated from a recent GWAS^7^. In the depression summary statistics, 358 (inclusive of 4 proxy variants with a minimum r^2^≥0.8) out of 403 independent (r^2^<0.01) type 2 diabetes SNPs were available for the analysis. To obtain the causal estimate, we applied the inverse variance weighted (IVW) method^5^. We performed sensitivity MR analysis using weighted median (WM)^10^, MR-Egger regression^11^, the simple mode^12^, and the weighted mode^12^ methods to evaluate the potential violations of the MR assumptions (**Supplementary Methods**) and confirm the robustness of the two-sample MR results from the IVW approach. F-statistic was used to evaluate the instrument strength, where F>10 indicates the presence of a strong instrument. The F-statistics indicated a good instrument strength for both type 2 diabetes (F-statistics = 61.26) and depression (F-statistics = 43).We assessed heterogeneity between the causal estimates from each SNP using Cochran’s Q-test. The sensitivity of causal inference to any individual genetic variant was tested by leave-one-out analysis. We used the STROBE-MR reporting guideline for MR studies to facilitate the readers’ evaluation of our results^13^ **(Supplementary Table 1)**.

### Genome-wide association studies (GWAS) and Multi-phenotype GWAS (MP-GWAS)

#### Cohorts used

##### 1) UK Biobank (UKBB)

We used data from the UK Biobank (UKBB, www.ukbiobank.ac.uk), which includes over 500,000 individuals from 22 centers across the United Kingdom. Study participants were between 40 and 69 years at recruitment and provided information including body measurements, biological samples, brain imaging data, socio-demographic and lifestyle indicators. Genetic data was available for 488,377 individuals in the UKBB genotyped using the UKBB BiLEVE array (n = 49,979) and the UKBB Axiom Array (n = 438,398)^14^. The genetic data was imputed using the Haplotype Reference Consortium^15^, the UK10K^16^ and 1000 Genomes Phase 3^17^, resulting in approximately 90 million variants available for association testing. This research has been conducted under application number 35327 and all participants gave informed consent during enrolment.

##### 2) FinnGen

We utilized the FinnGen summary statistics for replication of our single- and multi-phenotype association results (www.finngen.fi/fi). The June 2020 data freeze used in our analysis comprised of 135,638 individuals. Summary statistics for 1,801 phenotypes are publicly available for analysis. FinnGen study participants were genotyped using the Illumina and Affymetrix chip arrays (Illumina Inc., San Diego, and Thermo Fisher Scientific, Santa Clara, CA, USA, https://www.thermofisher.com/). The data was then imputed using the SISu v3 imputation panel (http://sisuproject.fi) resulting in 16,962,023 variants available for association analysis. FinnGen study participants were genotyped using the Illumina and Affymetrix chip arrays (Illumina Inc., San Diego, and Thermo Fisher Scientific, Santa Clara, CA, USA, https://www.thermofisher.com/). Summary statistics provided for the FinnGen data association analyses were generated using the SAIGE software^18^.

#### Phenotype definition

##### Type 2 diabetes

In the UKBB, type 2 diabetes cases were defined if individuals self-reported to have a diabetes diagnosis by a doctor, were on insulin medication one year after diagnosis, and were at least 40 years old at the time of diagnosis. Individuals not meeting these criteria were classified as controls. For both cases and controls, we excluded individuals with gestational diabetes (Field 4041, code = 1), those younger than 40 years at the time of diagnosis (Field 2976) and individuals on insulin medication within the first year of diagnosis (Field 2986). Sex discordant individuals (genotype vs. reported sex) were also excluded from the analyses. We restricted our analyses to European individuals to limit confounding by ancestry. In total, 19,344 cases and 463,641 controls were available (**Supplementary Table 2**).

The GWAS summary statistics in the FinnGen study were based on the ICD10-coded type 2 diabetes (ICD code E11) on 32,469 individuals (case/control numbers not available).

##### Depression

We defined depression in two ways: ICD-10 coded major depressive disorder (MDD) based on linked data from hospital records and self-reported depressive symptoms using the Patient Health Questionnaire 9 (PHQ-9)^19^.

##### ICD-coded MDD

Individuals with a primary diagnosis of a depressive episode (ICD code F32) and recurrent depression (ICD code F33) were defined as ICD-coded MDD cases (hereinafter referred to as MDD). Individuals who answered “NO” to the questions “Have you ever seen a general practitioner (GP) for nerves, anxiety, tension or depression?” (Field 2090) and “Have you ever seen a psychiatrist for nerves, anxiety, tension or depression?” and “NO” to either “depressed/down for a whole week” (Field 4598) or “Ever unenthusiastic/disinterested for a whole week” were set as controls. Participants were excluded from the study if they had a diagnosis of bipolar disorder (ICD codes F30, F31), mixed and other personality disorder (F61) and schizophrenia (ICD code F20). Participants on antipsychotic medication (Field 20003) for 58 drugs were also excluded. In total, 5,262 cases and 86,275 controls were used for the MDD phenotype (**Supplementary Table 1**). The proportion of MDD participants who had a type 2 diabetes diagnosis are shown in **Supplementary Table 3**.

FinnGen study data had the ICD-10 codes F32 and F33 available for MDD. The GWAS summary statistics on MDD were based on a total of 23,424 individuals (case/control numbers not available).

##### Depressive symptoms

In UKBB, self-reported depressive symptoms over the previous two weeks (from the time of study enrolment) were assessed using the (PHQ-9)^19^ questionnaire (**Supplementary Table 4**). It has been shown that the PHQ-9 questionnaire is invariant between people with and without diabetes^20^ suggesting its interpretation is similar for both diabetes cases and controls. Individuals missing responses for more than three PHQ-9 items were excluded from the analysis. Missing PHQ-9 responses for the remaining individuals were imputed using the ImputeSCOPA software (https://github.com/ImperialStatGen/imputeSCOPA), which implements a random forest approach to impute missing items. The variables sex, age, education qualification, body mass index (BMI), Townsend Deprivation Index^21^ (an area-based measure of deprivation), genotyping array and eight principal components (PCs) were included in the imputation model to improve the predictive accuracy of the imputation (**Supplementary Table 5**). The sum of all nine PHQ-9 items after imputation for everyone was used for quantitative association analysis. PHQ-9 data were available for 153,079 individuals (**Supplementary Table 1**). The proportion of individuals with PHQ-9 data and a type 2 diabetes diagnosis are shown in **Supplementary Tables 3 and 6**. Symptom-based depression phenotypes were unavailable in the FinnGen replication dataset.

##### GWAS

We performed separate GWAS for type 2 diabetes, MDD and PHQ-9 in UKBB data with BOLT-LMM using a linear mixed model^22^. We adjusted for age, sex, array, BMI and the first 8 PCs. We analyzed common variants (MAF>5%), with imputation scores >0.4, Hardy-Weinberg Equilibrium (HWE) p-value>1×10^−6^ and per SNP variant missingness<0.015. Manhattan plots were constructed using the *ggplot2* R package^23^. All analyses were performed on Human genome build 37. The statistical threshold for genome-wide significant SNPs (signals) used was *p*<5×10^−8^.

##### Multi-phenotype GWAS

We used MTAG (Multi-Trait Analysis of GWAS)^24^, which implements a generalized inverse-variance weighted meta-analysis, to increase the power for locus identification, improve SNP effect size estimates for type 2 diabetes and depressive phenotypes, and to identify potential multi-phenotype genetic variant effects. We used the summary statistics of the individual GWAS as inputs of MTAG. In UKBB, two MTAG models were tested - one with type 2 diabetes and MDD and another with type 2 diabetes and total PHQ-9 scores. This was to assess the consequence of using two different depression definition criteria (disease diagnosis vs. disease symptoms) in an MP-GWAS approach. In FinnGen, only the first approach was applied due to data availability. For each MTAG model tested, MTAG outputs phenotype specific association statistics. To assess the robustness of MP-GWAS results, MTAG computes a maximum false discovery rate (maxFDR) statistic, a theoretical upper bound limit on the FDR for a GWAS^24^. Lower maxFDR values (maxFDR<5%) indicate robust results.

### Expression quantitative trait *loci* (eQTL) analyses

To explore and identify target genes of the identified *loci*, we utilized several eQTL databases and datasets of relevant tissues. We extracted eQTL data from the GTEx Portal (https://gtexportal.org) for SNPs identified in our MP-GWAS, focusing on type 2 diabetes and depression relevant tissues (*i*.*e*., brain, muscle, liver). In addition, as GTEx does not include data from pancreatic islets, a crucial tissue in type 2 diabetes pathogenicity, we utilized recent eQTL data from Tiger T2D Systems (http://tiger.bsc.es) obtained from > 500 brain-dead organ donor islets^25^. We extracted data for the seven shared SNPs identified in our MP-GWAS from both eQTL studies (using nominal significance, *p*< 0.05).

Furthermore, we also used GTEx Version 7 transcriptome data^26^ from European individuals to identify eQTLs using our MP-GWAS summary statistics. We focused on relevant tissues in 1) type 2 diabetes, namely liver, whole pancreas, muscle, adipose subcutaneous, adrenal gland, whole blood, and 2) depression, including putamen basal ganglia, hippocampus, substantia nigra, frontal cortex, amygdala, anterior cingulate cortex. For each tissue, the predicted expression levels were then correlated with type 2 diabetes and PHQ-9 MTAG summary statistics. *P*-values were corrected for multiple testing using Bonferroni correction based on the number of genes tested per tissue (**Supplementary Table 7**). Genes where less than 80% of the SNPs used in the model were found in the GWAS summary statistics were excluded due to low reliability of association results. This analysis focused on type 2 diabetes and PHQ-9 phenotypes only as the MDD phenotype was underpowered in both GWAS and MP-GWAS.

## RESULTS

### Mendelian randomization

Our MR analysis revealed that depression was causally and positively associated with type 2 diabetes using the IVW method, with an OR of 1.18 (95%CI = 1.06-1.32; *p* = 0.0024). This result was consistent with the WM sensitivity analyses, which showed an OR of 1.11 (95%CI = 1.00-1.23, *p* = 0.043) (**Figure 1 and Supplementary Figure 2A, Supplementary Table 8**). The MR-Egger test showed no evidence of directional pleiotropy (*p* = 0.51), further confirming the validity of the results. Additionally, the leave-one-out analysis showed no outliers, suggesting that the observed association was not changed after removing any single variant (**Supplementary Figure 3**). The Cochran’s Q statistic for heterogeneity was significant for the IVW method (Q=261.62, *p*=1.26×10^−20^).

**Figure 1.**
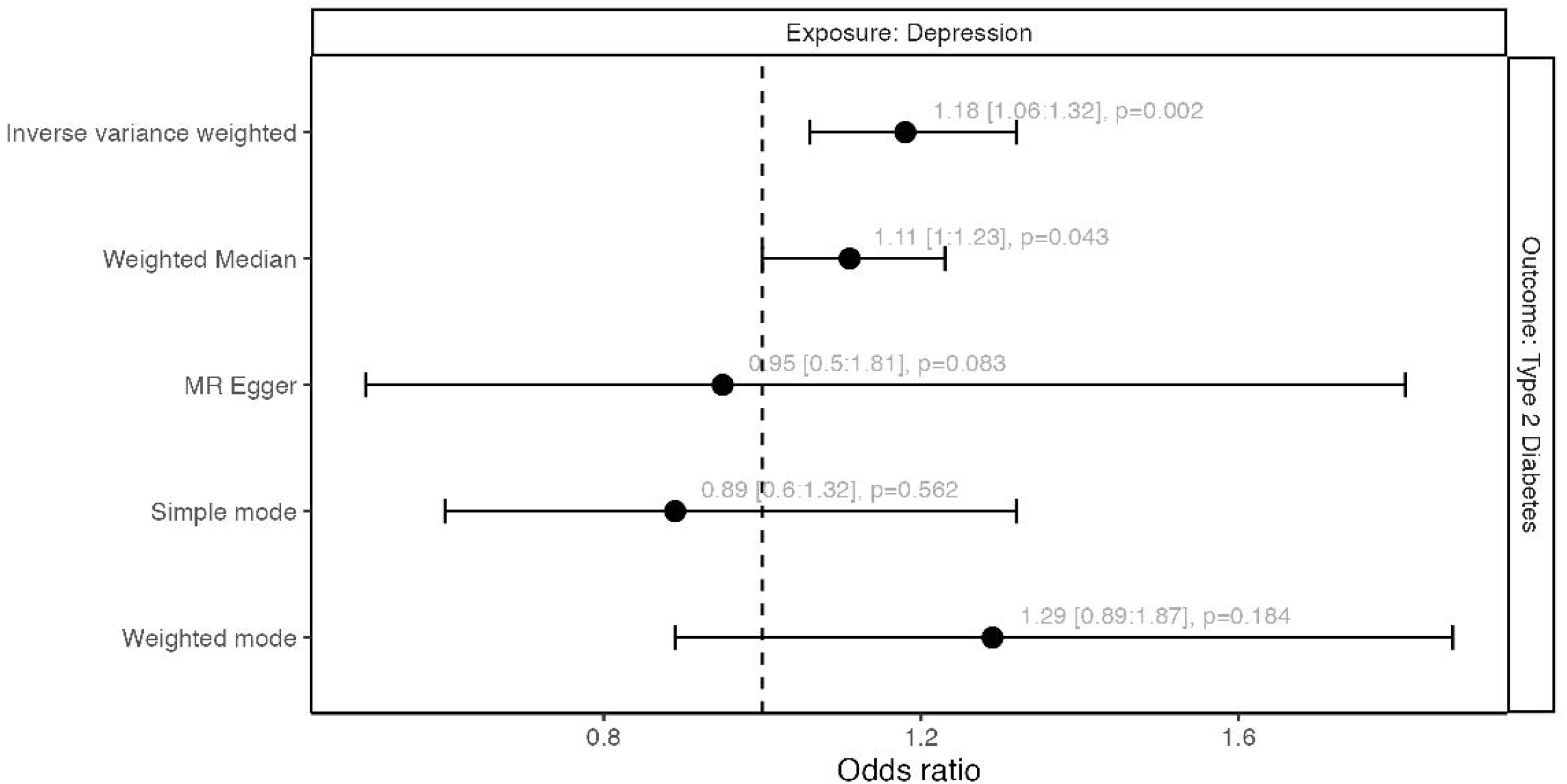
Forest plot showing the Mendelian randomization analysis results between depression (exposure) and type 2 diabetes (type 2 diabetes). The odds ratio (OR), their 95% confidence intervals and *P*-values are shown.

We found no evidence of causality in the reverse direction between type 2 diabetes and depression, in the primary nor sensitivity analysis (IVW: OR = 0.999; CI = 0.99-1.01; *p* = 0.843) (**Supplementary Figure 2B, Supplementary Table 9**).

### Genome wide association study in type 2 diabetes and depression

In order to identify whether type 2 diabetes and depression have a shared genetic etiology, we first performed a GWAS for both phenotypes, separately, using the UKBB. In total, for type 2 diabetes, we used 482,958 individuals (19,344 cases; 463,641 controls), and for depression we used 91,537 (5,262 cases; 86,276 controls) for MDD and 153,079 for PHQ-9. For type 2 diabetes, we identified 92 independent SNPs at 84 *loci*, of which 59 were replicated in the FinnGen with nominal significance (*p*<0.05) and consistent in direction of effect. For depression, we found three independent SNPs and *loci* for PHQ-9 and no SNPs associated with the binary depression MDD trait in the UKBB. None of the GWAS SNPs identified in type 2 diabetes were shared with depression (**Supplementary Figure 4, Supplementary Tables 10-12)**.

### Multi-phenotype GWAS in UKBB

To improve the power to identify shared genetic variants between the two phenotypes, we performed the largest-to-date MP-GWAS of type 2 diabetes and depressive phenotypes in the UKBB using the respective GWAS summary statistics.

The MP-GWAS model with type 2 diabetes and MDD did not identify any significant associations for the binary definition of depression (MDD). For type 2 diabetes, we identified 71 independent signals at 66 *loci* (**Supplementary Figure 5 and 6, Supplementary Table 10)**. The maxFDR for the type 2 diabetes and MDD MP-GWAS results was 1.5% and 7.7% respectively (**Supplementary Table 13**), indicating that the MDD results were likely inflated by the higher powered type 2 diabetes GWAS^24^. In FinnGen, the maxFDR were 11.5% and 25.5%, respectively, indicating highly inflated results for both traits.

In contrast, in the MP-GWAS model with type 2 diabetes and PHQ-9, we identified eight independent SNPs for PHQ-9 (compared to only three SNPs detected in GWAS) (**Figure 2, Supplementary Table 14**), suggesting greater power for SNP discovery. Only the *CACNA2D2* locus was reported in both GWAS and MP-GWAS analyses for PHQ-9, although with different, yet highly correlated lead SNPs (*CACNA2D2*, chromosome 2, SNP_SP-GWAS_ rs35335661, SNP_MP-GWAS_ rs1467916, r^2^=0.99). For type 2 diabetes, MP-GWAS identified 53 SNPs at 50 *loci* (compared to 92 identified in the single-phenotype GWAS), of which 24 *loci* were previously reported^7^. We replicated 37 type 2 diabetes signals in FinnGen cohort (**Supplementary Table 14**).

**Figure 2.**
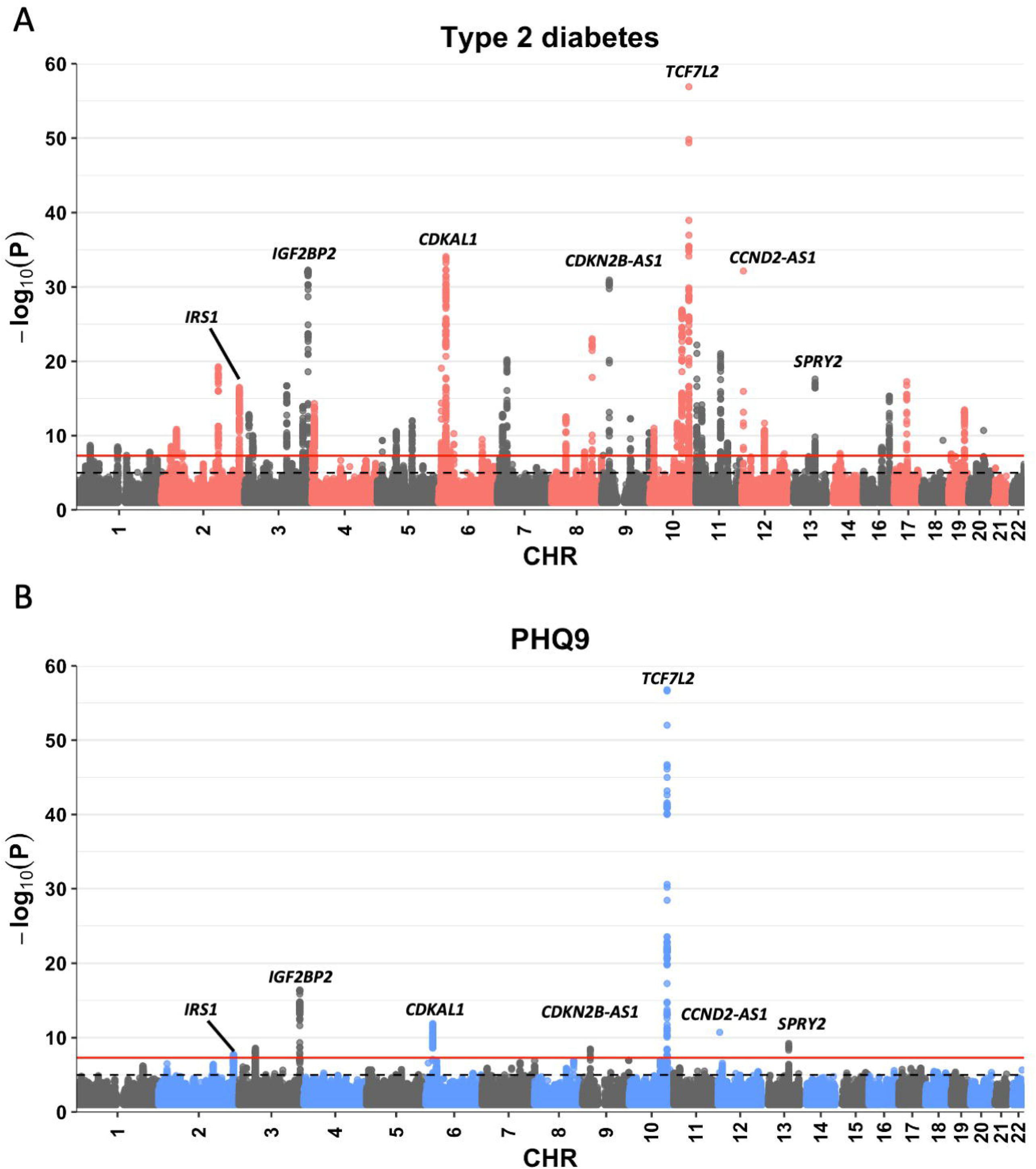
Manhattan plots of type 2 diabetes (A) and PHQ-9 (B) after MP-GWAS in UK Biobank. The red horizontal line shows genome-wide significance threshold (*P*<5×10^−8^). Grey dashed horizontal lines show suggestive genome-wide significance threshold (*P*<1×10^−5^). The shared loci are annotated.

In total, we found seven SNPs shared between type 2 diabetes and PHQ-9: rs7903146 (*TCF7L2*), rs7766070 (*CDKAL1*), rs1359790 (*SPRY2*), rs16860235 (*IGF2BP2*), rs76895963 (*CCND2-AS1*), rs2972144 (*IRS1*) and rs10811662 (*CDKN2B-AS1*) (**Table 1, Figure 2**). The maxFDR for type 2 diabetes and PHQ-9 after MP-GWAS were 0.98% and 1.8% respectively (**Supplementary Table 13**), indicating the results are robust and not influenced by the sample sizes of either GWAS.

**Table 1.** Summary statistics of the seven shared loci between type 2 diabetes and PHQ-9 after MP-GWAS.

|  |  |  |  |  |  |  | Type 2 diabetes |  |  | PHQ-9 |  |  |
| --- | --- | --- | --- | --- | --- | --- | --- | --- | --- | --- | --- | --- |
| CHR | SNP | BP | NEAREST<br>GENE | EA | NEA | EAF | BETA | SE | P | BETA | SE | P |
| 2 | rs2972144 | 227101411 | *IRS1* | G | A | 0.65 | 0.0031 | 0.00038 | 4.89x10<sup>-16</sup> | 0.065 | 0.012 | 1.80x10<sup>-08</sup> |
| 3 | rs16860235 | 185512361 | *IGF2BP2* | A | G | 0.28 | 0.0046 | 0.00040 | 1.41x10<sup>-30</sup> | 0.103 | 0.012 | 4.19x10<sup>-17</sup> |
| 6 | rs7766070 | 20686573 | *CDKAL1* * | A | C | 0.26 | 0.0046 | 0.00041 | 7.79x10<sup>-29</sup> | 0.103 | 0.014 | 1.24x10<sup>-12</sup> |
| 9 | rs10811662 | 22134253 | *CDKN2B-AS1* | G | A | 0.83 | 0.0049 | 0.00048 | 7.88x10<sup>-25</sup> | 0.09 | 0.015 | 4.30x10<sup>-09</sup> |
| 10 | rs7903146 | 114758349 | *TCF7L2* | T | C | 0.29 | 0.011 | 0.00040 | 6.88x10<sup>-178</sup> | 0.21 | 0.012 | 1.68x10<sup>-64</sup> |
| 12 | rs76895963 | 4384844 | *CCND2-AS1* | T | G | 0.98 | 0.015 | 0.0014 | 3.08x10<sup>-27</sup> | 0.29 | 0.043 | 1.91x10<sup>-11</sup> |
| 13 | rs1359790 | 80717156 | *SPRY2* | G | A | 0.72 | 0.0035 | 0.00040 | 8.54x10<sup>-17</sup> | 0.076 | 0.012 | 5.91x10<sup>-10</sup> |

\* Different SNP for PHQ-9 (rs2206734, Position-20694884, Effect Allele-G, Non-effect
Allele-C; In high LD with type 2 diabetes SNP=R2=0.544)
Legend: CHR=chromosome, BP=base pair position (genome build 37), EA=effect allele, NEA=non-effect allele, EAF=effect allele frequency

### eQTL analyses

To explore whether the seven SNPs shared between type 2 diabetes and self-reported depression had a downstream functional impact, we first extracted eQTL data of the seven identified SNPs using the 1) GTEx Portal in relevant tissues, including muscle, liver and brain, and 2) the Tiger Portal for pancreatic islets. We found that of the seven shared SNPs between type 2 diabetes and depression, six SNPs were associated with the expression of nearby genes in relevant tissues (**Table 2**). This includes rs2972144-G risk allele associated with a decreased expression of *IRS1* and *RP11-395N3*.*2* in visceral and subcutaneous adipose and *RP11-395N3*.*1* in subcutaneous adipose tissue. Additionally, the rs76895963-T risk allele was associated with the decreased expression of *CCND2* in pancreatic islets, brain cerebellum, skeletal muscle and subcutaneous adipose tissue, the decreased expression of its antisense *CCND2-AS1* in pancreatic islets, brain cerebellum, basal ganglia and cortex, in addition to *CCND2-AS2* in the cerebellum. For pancreatic islets, we found an increased expression of *NDUFA9* (**Table 2**).

**Table 2.**
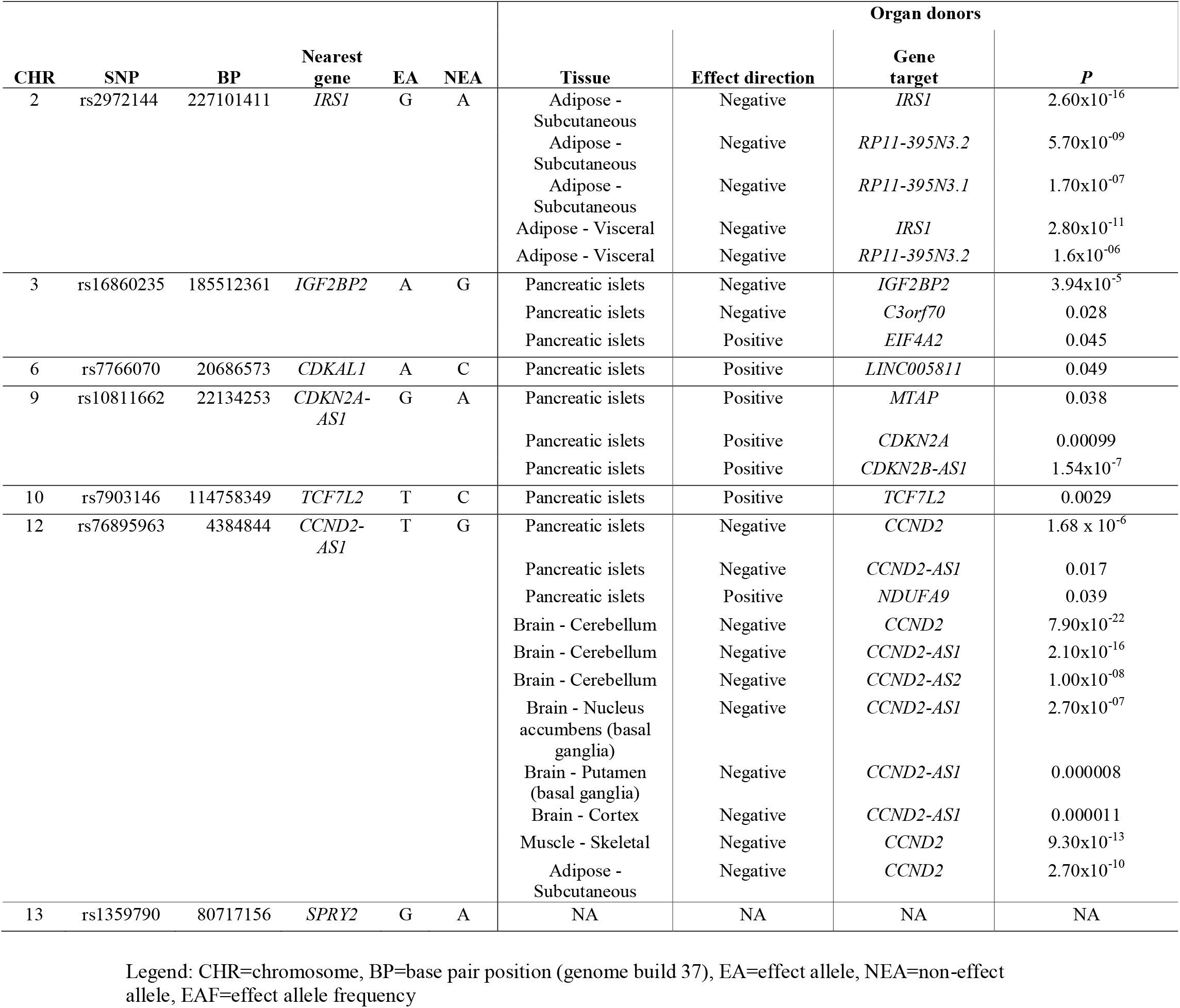
Target genes of SNPs shared type 2 diabetes and depressive symptoms in relevant target tissues.

In addition, using our MP-GWAS summary statistics, we tested in GTEx whether type 2 diabetes and PHQ-9 were associated with shared gene expression changes in several target tissues except for pancreatic islets as GTEx does not include data on that. This analysis identified additional target genes associated with both type 2 diabetes and PHQ-9 in eight tissues, consistent in direction of effect for both phenotypes: adipose subcutaneous (*IRS1, NCR3LG1, RP11-395N3*.*2*), amygdala (*HSPA1B*), frontal cortex (*CDKAL1*), hypothalamus (*EIF2S2P3*), skeletal muscle (*HLA-DRA, RP11-370C19*.*2*), substantia nigra (*BETL1*), and whole blood (*EIF2S2P3, HLA-DRB1*) (**Supplementary Figure 7, Supplementary Table 7 and 15-17**). Altogether, our data demonstrates a functional impact of our *loci* in several related tissues.

## CONCLUSIONS

We performed a large comprehensive study to investigate the relationship between type 2 diabetes and depression and found evidence for a causal positive association from depression to type 2 diabetes. We also performed a multi-phenotype GWAS of the two diseases, highlighting seven shared *loci* that target nearby genes in several target tissues. Altogether, our study provides novel insight into the underlying mechanisms linking the two diseases.

Our MR analysis, based on recent large-scale GWAS, provides evidence for causality on the previously reported epidemiological associations from depression to type 2 diabetes^3^, but not in the reverse direction. These findings are consistent with the pathophysiology for these diseases, whereby: 1) depression starts in adolescence or early adulthood^27^, whereas type 2 diabetes usually develops later^28^, 2) poor health habits among individuals with depression, including smoking, physical inactivity and increased caloric intake (and associated overweight), that are known to facilitate the development of type 2 diabetes^3,29^, 3) antidepressants frequently induce weight gain leading to type 2 diabetes ^29^, and 4) the systemic inflammation associated to increased stress hormone levels such as cortisol in the context of depression also favors insulin resistance^30^.

While epidemiological studies also show increased risk of depression in people with type 2 diabetes^4^, we did not find evidence for causality in this relationship. Our evaluation of instrument strength for the MR analysis suggests that the lack of this causal association is not simply an issue of statistical power. A likely hypothesis is that the epidemiologically observed association is confounded via other factors, which are not easily assessed in epidemiological studies, such as psychosocial factors related to the painful management of a middle-age chronic disease. Diabetes distress, the psychological burden caused by dealing with having diabetes and having to care for it, has indeed been linked to depression^31^. However, a large international longitudinal study of 14 countries has shown that only depressive symptoms rather than MDD were predicted by diabetes distress one year after diagnosis^32^.

In addition, our multi-phenotype GWAS revealed seven shared SNPs between type 2 diabetes and self-reported depressive symptoms consistent in their directions of effect. It is interesting to note that we did not find any shared SNPs associated with the binary MDD clinical diagnosis and type 2 diabetes which we speculate could be due to several reasons: 1) that the association between type 2 diabetes and depression is due to the less strictly defined symptom-based depressive measures, and 2) the PHQ-9 is a continuous definition of depression, and likely improves power compared to the binary MDD definition. Indeed, previous studies showed that depression defined based on self-reported symptoms (minimal phenotyping) rather than strict diagnostic criteria enables greater power for locus discovery in GWAS^33^.

Our eQTL analysis provided some clues to the underlying mechanisms linking the two diseases. For instance, we found that the rs76895963-T risk allele was associated with the decreased expression of *CCND2* in the brain and pancreatic islets and insulin target tissues (adipose and skeletal muscle), suggesting a pleiotropic effect of this *locus. CCND2* encodes Cyclin D2, which is involved in cell cycle regulation and with a role in pancreatic beta cell proliferation and insulin secretion^34^, consistent with our eQTL data showing a decreased expression of the gene in pancreatic islets. Mutations in *CCND2* have been described in individuals with brain malformations^35^, confirming a role in maintaining brain growth, and mouse knockout models of *CCND2* have no brain neurogenesis and showed mild depression-like symptoms that were alleviated by anti-depressant chronic fluoxetine treatment^36^. In addition, a recent study showed that neurogenesis in the brain may prevent depressive symptoms. Indeed, these studies demonstrate a potential mechanism whereby *CCND2* down-regulation may halt this process^37^. In addition, the inhibition of *CCND2* in adipose tissue has been shown to dysregulate adipocyte differentiation^38^ and downregulated in obesity, suggesting a key role in maintaining adipocyte regeneration^39^. Therefore, we show that *CCND2* has a multi-system and pleiotropic effect and could potentially mediate this relationship between type 2 diabetes and depression.

In addition, we found that the rs2972144-G risk allele was associated with decreased expression of *IRS1*, which encodes insulin receptor substrate 1, in adipose tissue. *IRS1* is a key signaling molecule necessary for insulin response in insulin target tissues and has been shown to be associated with insulin resistance^40^, a condition linked to both the development of type 2 diabetes and depression^41^.

Also, our eQTL analyses revealed further insights into the potential mechanisms underlying the relationship between type 2 diabetes and depression. Lower expression of *CDKAL1* in frontal cortex was associated both with type 2 diabetes and depressive symptoms. Although reported for bipolar disorder^42,43^, variation in *CDKAL1* has been associated with depressive phenotypes only in one previous study^44^. In addition, although we did not find an eQTL for this *locus* in pancreatic islets in our study, variation at *CDKAL1* has been implicated in type 2 diabetes and shown to reduce insulin secretion^45^.

We also found two target genes implicating the HLA region (*HLA-DRA, HLA-DRB1*) in the shared pathogenesis of the two diseases, in blood and skeletal muscle. The role of depression treatments, targeting the immune system, is very active^46^. Type 2 diabetes is also known to have an impact on immune system with high blood glucose levels causing an inflammatory response^47^. While trying to underpin the molecular mechanisms within this potential shared pathway, further studies on the type 2 diabetes-depression comorbidity should account also for the role of obesity with its known links with inflammation^48^, type 2 diabetes^49^ and suggested with depression^50^.

Our results highlight a shared genetic effect between the two phenotypes with plausible biological significance that explain how environmental factors (*i*.*e*., stress, lifestyle habits and anti-depressant medication) could lead to the underlying co-morbidity. Therefore, we speculate that our results could hold some clinical significance. For instance, the choice of anti-depressant treatment offered to people with depression at risk of type 2 diabetes should favor those that provide better glycemic control such as selective serotonin reuptake inhibitors (SSRIs)^51^. Additionally, people with depression should be encouraged, as part of routine clinical care, to promote positive lifestyle habits such as increased physical activity, adequate sleep, and a proper dietary regime.

This study exhibits several strengths. We report the first study to investigate the causal relationship of type 2 diabetes and depression in both directions and performed the largest-to-date MP-GWAS for the two diseases. Our investigation highlights the ability of multi-phenotype approaches to reveal the shared associations in co-morbid diseases and further confirms the power of large-scale datasets to uncover phenotype associations, including self-reported and continuous measures of diseases and their symptoms. However, there are some limitations to be considered. Our main UKBB analyses on MDD did not yield any genome-wide significant signals in both GWAS and MP-GWAS and a validation in larger datasets is needed. In addition, to effectively probe the credibility of the MTAG MP-GWAS results, replication using another MP-GWAS method is needed as part of future analyses. Finally, we could only replicate the type 2 diabetes-MDD analyses due to unavailability of publicly available depressive symptoms GWAS and in the FinnGen cohort.

In conclusion, the shared *loci* between depressive symptoms and type 2 diabetes support a pleiotropic role in target tissues, providing insight into their pathophysiology and co-morbidity, boosting our understanding of the pathogenesis of these two diseases. Additionally, self-reported depression/depressive symptoms may offer more in deciphering the underlying co-morbidity with type 2 diabetes compared to the strictly defined MDD. The causal effect of depression leading to the development of type 2 diabetes has important implications for a more efficient prevention of type 2 diabetes from early adulthood.

## Data Availability

All data produced in the present study are available upon reasonable request to the authors

## ACKNOWLEDGEMENTS

This research has been conducted using the UK Biobank Resource under Application Number 35327. This research was in part funded by the Diabetes UK (BDA number: 20/0006307), the European Union’s Horizon 2020 research and innovation programme (LONGITOOLS, H2020-SC1-2019-874739), Agence Nationale de la Recherche (PreciDIAB, ANR-18-IBHU-0001), by the European Union through the “Fonds européen de développement regional” (FEDER), by the “Conseil Régional des Hauts-de-France” (Hauts-de-France Regional Council) and by the “Métropole Européenne de Lille” (MEL, European Metropolis of Lille).

