## Supplementary for "Bi-directional Mendelian randomization and multi-phenotype GWAS show causality and shared pathophysiology between depression and type 2 diabetes"

**Supplementary methods**

**UK Biobank**

In the UK Biobank (UKBB), electronic health records from the National Health Service registers are linked to the resource and enable follow-up of participants using ICD-9 and ICD-10 (International Classification of Diseases, ninth and tenth versions, respectively) codes. This, in addition to self-reported disease status at the time of recruitment, enables researchers define various disease phenotypes. Hospital admission records are linked to the UKBB through the data fields 41202 and 41204 representing primary and secondary disease diagnosis, respectively.

**FinnGen**

FinnGen study participants were genotyped using the Illumina and Affymetrix chip arrays (Illumina Inc., San Diego, and Thermo Fisher Scientific, Santa Clara, CA, USA, https://www.thermofisher.com/). The data was then imputed using the SISu v3 imputation panel (http://sisuproject.fi) resulting in 16,962,023 variants available for association analysis. Summary statistics provided for the FinnGen data association analyses were generated using the SAIGE software^1^.

**Multi-Trait Analyses of GWAS (MTAG)**

MTAG performs an inverse-variance weighted meta-analyses of related phenotypes (ref). MTAG uses GWAS summary statistics (effect size estimates and their standard errors (BETA, SE) and the *P*-values of the associations). Formatting of the summary statistics was done as recommended by the authors^2^. To account for sample overlap present in our UKBB analyses, MTAG applies the bivariate linkage disequilibrium (LD) score regression to estimate the correlation in GWAS estimation error due to sample overlap.

**Mendelian randomization**

Causality can be established in MR only if it uses valid instrumental variables (IV) defined by three core assumptions: (1) that they have a true effect on the exposure (the relevance assumption) (2) that they only affect the outcome through the risk factor (the exclusion restriction assumption) (3) that they are independent of any measured and unmeasured confounding factors of the exposure–outcome relationship (the independence assumption)^3^. The genetic instruments for depression were from a recent large-scale GWAS of depression that reported 102 independent association signals^4^. They meta-analyzed data for 807,553 individuals, including 246,363 from UK Biobank, 23andMe and Psychiatric Genomics Consortium. The cases in the tree contributing GWASs were defined using self-reported and clinical data^4^.

### **LD clumping**

To identify independent loci and their lead SNPs for each association result (SP-GWAS and MP-GWAS), we applied a LD-based clumping algorithm performed in PLINK^5^. The following parameters were set: *-clump-p1* 5×10^-8^ (significance threshold for the index SNP), *-clump-p2* 1×10^-5^ (significance threshold for the clumped SNPs), *-clump-r^2^* 0.01 (LD threshold for clumping) and *–clump-kb* 500kb (distance threshold for clumping). Clumped signals were annotated to their nearest gene using the ANNOVAR software^6^.

**List of supplementary tables**

**Supplementary Table 1**. STROBE-MR checklist for Mendelian Randomization analysis.

**Supplementary Table 2**. UKBB study participants, including gender proportions, used in the present study

**Supplementary Table 3.** Proportion of depression phenotype participants with a type 2 diabetes diagnosis in the UK Biobank. Only individuals with both data available, depression and type 2 diabetes, are included

**Supplementary Table 4.** The PHQ-9 questionnaire as provided in the UK Biobank

**Supplementary Table 5.** The list of variables used in the imputation model for missing PHQ-9 items. The data to be imputed included only individuals with less than three PHQ-9 item responses missing

**Supplementary Table 6.** Proportion of type 2 diabetes cases and controls among depression severity categories defined by the PHQ-9 sum scores

**Supplementary Table 7.** The number of genes tested per GTEx tissue model and the related significance threshold, followed by the number of significant (Bonferroni corrected) genes for Type 2 diabetes and PHQ-9 after MP-GWAS and finally, the number of common genes between type 2 diabetes and PHQ-9

**Supplementary Table 8.** Mendelian Randomization results with type 2 diabetes as the outcome and depression as the exposure variable

**Supplementary Table 9.** Mendelian Randomization results with Depression as the outcome and Type 2 Diabetes as the exposure variable

**Supplementary Table 10.** The number of genome-wide significant independent signals for each studied phenotype, identified with each analytical approach, implemented in either BOLT-LMM or MTAG in UKBB

**Supplementary Table 11.** Summary statistics for signals reaching genome-wide significance in GWAS of PHQ9 and type 2 diabetes in UKBB and their replication in FinnGen

**Supplementary Table 12**. Signals reaching suggestive evidence of association (*P*<10-6) in GWAS of MDD in UKBB and their replication in FinnGen

**Supplementary Table 13**. maxFDR for MTAG results in each model analyzed

**Supplementary Table 14**. Signals reaching genome-wide significance for type 2 diabetes in the MP-GWAS of PHQ-9 and type 2 diabetes

**Supplementary Table 15**. Table showing type 2 diabetes and PHQ9 significant eQTL genes after MP-GWAS and common genes between the two phenotypes per tissue

**Supplementary Table 16.** Table showing type 2 diabetes MetaXcan eQTL results for the significant genes in the tissues analyzed after MP-GWAS with PHQ-9

**Supplementary Table 17**. Table showing PHQ9 MetaXcan eQTL results for the significant genes in the tissues analyzed after MP-GWAS with type 2 diabetes

**List of supplementary figures**

**Supplementary Figure 1**. Mendelian Randomization analysis results between depression and type 2 diabetes

**Supplementary Figure 2**. A) Scatter plot for MR analyses of the causal effect of Depression on Type 2 Diabetes. B) Scatter plot for MR analyses of the causal effect of Type 2 Diabetes on Depression.

**Supplementary Figure 3**. Leave-one-out analysis: each row represents a MR analysis of Depression on Type 2 Diabetes

**Supplementary Figure 4**. Manhattan plots for type 2 diabetes, PHQ-9 and MDD GWAS in the UK Biobank

**Supplementary Figure 5**. Manhattan plots for type 2 diabetes and MDD after MP-GWAS in MTAG

**Supplementary Figure 6**. Manhattan plots for type 2 diabetes and MDD after MP-GWAS in FinnGen dataset

**Supplementary Figure 7.** Volcano plots of association in tissues implicated in both type 2 diabetes and PHQ-9

**Supplementary table 1**. STROBE MR checklist for Mendelian Randomization analysis.

| **Item** | **Complete/location** |
| --- | --- |
| 1. Title and Abstract: "Mendelian randomization" is named in the title or in the abstract | "Mendelian randomization" is named in the title and abstract |
| **Introduction** |  |
| 2. Background: Explain the scientific background and rationale for the reported study. Is causality between exposure and outcome plausible? Justify why MR is a helpful method to address the study question. | In the Introduction, we discuss why type 2 diabetes (T2D) is a plausible causal exposure for depressive symptoms and vice versa. In the first paragraph of Introduction section, we discuss why Mendelian randomization is useful in testing causality between depressive symptoms and type 2 diabetes. |
| 3. Objectives: State specific objectives clearly, including pre-specified causal hypotheses (if any). | In the second paragraph of Introduction, we describe the causal hypotheses of our study. |
| **Methods** |  |
| 4. Study design and data sources: Present key elements of study design early in the paper. Consider including a table listing sources of data for all phases of the study. For each data source contributing to the analysis, describe the following:   a) Describe the study design and the underlying population from which it was drawn. Describe also the setting, locations, and relevant dates, including periods of recruitment, exposure, follow-up, and data collection, if available.  b) Give the eligibility criteria, and the sources and methods of selection of participants.  c) Explain how the analyzed sample size was arrived at.  d) Describe measurement, quality and selection of genetic variants.  e) For each exposure, outcome and other relevant variables, describe methods of assessment and, in the case of diseases, the diagnostic criteria used.  f) Provide details of ethics committee approval and participant informed consent, if relevant. | Available information about the GWAS studies is provided in the "Mendelian Randomization" section of the “Research design and Methods” section. Further information is given in the original GWAS publications referenced. |
| 5. Assumptions: Explicitly state assumptions for the main analysis (e.g. relevance, exclusion, independence, homogeneity) as well assumptions for any additional or sensitivity analysis. | Stated in the "Mendelian Randomization" section of the “Research design and Methods” section for both the main and the sensitivity analyses. |
| 6. Statistical methods main analysis   Describe statistical methods and statistics used.  a) Describe how quantitative variables were handled in the analyses (i.e., scale, units, model).  b) Describe the process for identifying genetic variants and weights to be included in the  analyses (i.e., independence and model). Consider a flow diagram.  c) Describe the MR estimator, e.g. two-stage least squares, Wald ratio, and related statistics.  Detail the included covariates and, in case of two-sample MR, whether the same covariate set was used for adjustment in the two samples.  d) Explain how missing data were addressed.  e) If applicable, say how multiple testing was dealt with. | a) we used diagnosis of T2D and depression as binary phenotypes  b) described in the "Mendelian Randomization" section of the “Research design and Methods” section  c) age, sex and principal components were used as covariates  d) We stated that independent SNPs were used and described the proxy selection in the "Mendelian Randomization" section of the “Research design and Methods” section  e) N/A |
| Assessment of assumptions: Describe any methods used to assess the assumptions or justify their validity. | Our test for instrument heterogeneity and horizontal pleiotropy are described in the "Mendelian Randomization" section of the “Research design and Methods” section. |
| 8. Sensitivity analyses: Describe any sensitivity analyses or additional analyses performed. | We applied the weighted median (WM), MR-Egger regression, the simple mode and the weighted mode methods as sensitivity analysis as described in the "Mendelian Randomization" section of the “Research design and Methods” section. |
| 9. Software and pre-registration   a) Name statistical software and package(s), including version and settings used.  b) State whether the study protocol and details were pre-registered (as well as when and where). | a) All statistical software used are described in the "Mendelian Randomization" section of the “Research design and Methods” section  b) The study protocol was not pre-registered. |
| **Results** |  |
| 10. Descriptive data   a) Report the numbers of individuals at each stage of included studies and reasons for exclusion. Consider use of a flow-diagram.  b) Report summary statistics for phenotypic exposure(s), outcome(s) and other relevant variables (e.g. means, standard deviations, proportions).  c) If the data sources include meta-analyses of previous studies, provide the number of studies, their reported ancestry, if available, and assessments of heterogeneity across these studies. Consider using a supplementary table for each data source.  d) For two-sample Mendelian randomization:  i. Provide information on the similarity of the genetic variant-exposure associations between the exposure and outcome samples.  ii. Provide information on extent of sample overlap between the exposure and outcome data sources. | a) Information is given in the "Mendelian Randomization" section of the “Research design and Methods” section.  b) We give the summary statistics for our instruments in "Mendelian Randomization" section of the “Research design and Methods” section.  c) Provided in the Supplementary figure 1  d) We stated that samples are non-overlapping in the "Mendelian Randomization" section of the “Research design and Methods” section |
| 11. Main results   a) Report the associations between genetic variant and exposure, and between genetic variant and outcome, preferably on an interpretable scale (e.g. comparing 25th and 75th percentile of allele count or genetic risk score, if individual-level data available).  b) Report causal effect estimate between exposure and outcome, and the measures of uncertainty from the MR analysis. Use an intuitive scale, such as odds ratio, or relative  risk, per standard deviation difference.  c) If relevant, consider translating estimates of relative risk into absolute risk for a meaningful time-period.  d) Consider any plots to visualize results (e.g. forest plot, scatterplot of associations between genetic variants and outcome versus between genetic variants and exposure). | We describe the causal estimates in the "Mendelian Randomization" section of the “Results” section, 95% confidence intervals are provided as measures of uncertainty. Also two scatterplots included in the Supplementary Figure 3. |
| 12. Assessment of assumptions   a) Assess the validity of the assumptions.  b) Report any additional statistics (e.g., assessments of heterogeneity, such as I2, Q statistic). | Our test for instrument heterogeneity and horizontal pleiotropy are described in the "Mendelian Randomization" section of the “Research design and Methods” and “Results” section. |
| 13. Sensitivity and additional analyses   a) Use sensitivity analyses to assess the robustness of the main results to violations of the assumptions.  b) Report results from other sensitivity analyses (e.g., replication study with different dataset, analyses of subgroups, validation of instrument(s), simulations, etc.).  c) Report any assessment of direction of causality (e.g., bidirectional MR).  d) When relevant, report and compare with estimates from non-MR analyses.  e) Consider any additional plots to visualize results (e.g., leave-one-out analyses). | We report the results of all additional and sensitivity analyses together with the main results in the "Mendelian Randomization" section of the “Results” section.  c) We assessed causality in both directions.  d) Comparison with the epidemiological studies was reported in the Abstract, Introduction and Conclusion.  e) Results of leave-one-out analysis presented on the Supplementary Figure 4. |
| **Discussion** |  |
| 14. Key results | We discuss key results in the Conclusion section. “We conclude that the shared loci between depressive symptoms and type 2 diabetes support a role of insulin function and signaling and, immune system pathways in their pathophysiology and co-morbidity, boosting our understanding of the pathogenesis of the two diseases. The causal effect of depression leading to the development of type 2 diabetes is an important clinical management avenue for healthcare and prevention.” |
| 15. Limitations   Discuss limitations of the study, taking into account the validity of the MR assumptions, other sources of potential bias, and imprecision. Discuss both direction and magnitude of any potential bias, and any efforts to address them. | Described in the Conclusion section. |
| 16. Interpretations   a) Give a cautious overall interpretation of results considering objectives and limitations.  Compare with results from other relevant studies.  b) Discuss underlying biological mechanisms that could be modelled by using the genetic  variants to assess the relationship between the exposure and the outcome.  c) Discuss whether the results have clinical or policy relevance, and whether interventions  could have the same size effect. | We interpret the results in the abstract “Our findings support a causal effect from depression to type 2 diabetes, provide evidence for shared genetic background, and point to pathways related to insulin signaling and immune response in this co-morbidity” as well as in the Conclusion. |
| 17. Generalizability: | In the abstract we concluded “The causal effect of depression leading to the development of type 2 diabetes is an important clinical management avenue for healthcare and prevention”. |
| 18. Funding: | We have reported all sources of funding. |
| 19. Data and data sharing: | We give access information to all data used in the study in the "Data availability" section and our Supplementary tables 1-3. Links to the statistical software used, including TwoSampleMR, LD score regression, are given in the “Genetic correlation analysis”, “MR analysis” sections of Methods. |
| 20. Conflicts of Interest: | All authors have declared conflicts of interest (none reported). |

**Supplementary Table 2**. UKBB study participants, including gender proportions, used in the present study.

| Phenotype | Cases  % (male, female) | Controls  % (male, female) | Prevalence  % | Total  (*N*) |
| --- | --- | --- | --- | --- |
| Type 2 diabetes | 19,344  (64.3, 35.7) | 463,641  (45.0, 55.0) | 4.2 | 482,985 |
| MDD | 5,262  (32.6, 67.4) | 86,275  (47.3, 52.7) | 6.2 | 91,537 |
| PHQ-9 | 153,079  (43.5, 56.5) | - | - | 153,079 |

| Depression  Phenotype | Type 2 diabetes  Cases (%) | Type 2 diabetes  Controls (%) | Total | X^2^  statistic | X^2^  *P*-value |
| --- | --- | --- | --- | --- | --- |
| MDD Cases | 57 (1.10) | 5,283 (98.90) | 5,340 | 134.09 | <2.2x10^-16^ |
| MDD Controls | 3,756 (4.38) | 82,009 (94.62) | 85,767 |  |  |
| PHQ-9  (Sum score ≥ 10) | 2,781 (2.70) | 100,029 (97.30) | 102,810 | 22.41 | 2.78x10^-07^ |
| PHQ-9  (Sum score <10) | 1,116 (2.26) | 48,253 (97.74) | 49,369 |  |  |

**Supplementary Table 3.** Proportion of depression phenotype participants with a type 2 diabetes diagnosis in the UK Biobank. Chi-square test was done to test independence between groups.

| **Supplementary Table 4.** The PHQ-9 questionnaire as provided in the UK Biobank. | | |
| --- | --- | --- |
| UK Biobank Field | Field Description | Code |
| 20514 | Recent lack of interest or pleasure in doing things | 0,1,2,3 |
| 20510 | Recent feelings of depression | 0,1,2,3 |
| 20517 | Trouble falling or staying asleep or sleeping too much | 0,1,2,3 |
| 20519 | Recent feelings of tiredness or low energy | 0,1,2,3 |
| 20511 | Recent poor appetite or overeating | 0,1,2,3 |
| 20507 | Recent feelings of inadequacy | 0,1,2,3 |
| 20508 | Trouble concentrating on things | 0,1,2,3 |
| 20518 | Recent change in speed/amount of moving or speed | 0,1,2,3 |
| 20513 | Recent thoughts of suicide or self-harm | 0,1,2,3 |
| Code legend: 0 = Not at all, 1 = Several days, 2 = Half the days, 3 = Nearly every day | |  |

| **Supplementary Table 5.** List of variables in the data used in PHQ-9 items imputation. Data included only individuals with less than three PHQ9 items responses missing. | | | |
| --- | --- | --- | --- |
| **Variables** | **Missing Items** | **Missingness rate (%)** | **Items present** |
| Sex | 0 | 0 | 153 079 |
| Age | 0 | 0 | 153 079 |
| Array | 0 | 0 | 153 079 |
| Education qualification | 355 | 0.23 | 152 724 |
| Townsend Deprivation Index | 191 | 0.12 | 152 888 |
| BMI | 326 | 0.21 | 152 753 |
| PC1 | 0 | 0 | 153 079 |
| PC2 | 0 | 0 | 153 079 |
| PC3 | 0 | 0 | 153 079 |
| PC4 | 0 | 0 | 153 079 |
| PC5 | 0 | 0 | 153 079 |
| PC6 | 0 | 0 | 153 079 |
| PC7 | 0 | 0 | 153 079 |
| PC8 | 0 | 0 | 153 079 |
| phq9_1 | 306 | 0.20 | 152 773 |
| phq9_2 | 426 | 0.28 | 152 653 |
| phq9_3 | 238 | 0.16 | 152 841 |
| phq9_4 | 244 | 0.16 | 152 835 |
| phq9_5 | 146 | 0.095 | 152 933 |
| phq9_6 | 658 | 0.43 | 152 421 |
| phq9_7 | 143 | 0.093 | 152 936 |
| phq9_8 | 210 | 0.14 | 152 869 |
| phq9_9 | 1 118 | 0.73 | 151 961 |

| **Supplementary Table 6.** Proportion of type 2 diabetes cases and controls by depression severity categories defined by the PHQ-9 sum scores. | | | |
| --- | --- | --- | --- |
| Depression severity (PHQ-9 Sum scores) | Type 2 diabetes  Cases (%) | Type 2 diabetes  Controls (%) | Total |
| None - minimal (0 - 4) | 2,820 (2.34) | 117,515 (97.66) | 120,335 |
| Mild (5 - 9) | 690 (3.03) | 22,082 (96.97) | 22,772 |
| Moderate (10 - 14) | 251 (4.26) | 5,640 (95.74) | 5,891 |
| Moderately severe (15 - 19) | 92 (4.33) | 2,031 (95.67) | 2,123 |
| Severe (20 - 27) | 37 (4.32) | 819 (95.68) | 856 |
| Total | 3,890 | 148,087 | 151,977 |
| Only individuals with both PHQ-9 data and a type 2 diabetes diagnosis (N = 152,354) are included. | | | |

| **Supplementary Table 7.** The number of genes tested per GTEx tissue model and the related significance threshold, followed by the number of significant (Bonferroni corrected) genes for Type 2 diabetes and PHQ-9 after MP-GWAS and finally, the number of common genes between type 2 diabetes and PHQ-9. | | | | | |
| --- | --- | --- | --- | --- | --- |
| **Tissue** | **Genes**  **tested** | **Significance**  **threshold** | **No of significant**  **type 2 diabetes**  **genes** | **No of significant**  **PHQ9 genes** | **Common**  **Significant genes** |
| Adipose subcutaneous | 8,258 | 6.050x-06 | 27 | 4 | 3 |
| Adrenal gland | 4,602 | 1.086x-05 | 4 | 0 | 0 |
| Amygdala | 2,358 | 2.120x-05 | 4 | 2 | 1 |
| Anterior cingulate cortex | 3,302 | 1.514x-05 | 1 | 0 | 0 |
| Frontal cortex | 3,598 | 1.390x-05 | 5 | 1 | 1 |
| Hippocampus | 2,816 | 1.776x-05 | 5 | 0 | 0 |
| Hypothalamus | 2,832 | 1.766x-05 | 2 | 2 | 1 |
| Liver | 3,345 | 1.495x-05 | 7 | 0 | 0 |
| Pancreas | 5,334 | 9.370x-06 | 11 | 0 | 0 |
| Putamen basal ganglia | 3,174 | 1.575x-05 | 2 | 0 | 0 |
| Skeletal muscle | 7,514 | 6.560x-06 | 11 | 3 | 2 |
| Substantia nigra | 2,041 | 2.450x-05 | 1 | 1 | 1 |
| Whole blood | 6,285 | 7.960x-06 | 10 | 2 | 2 |
| Significance threshold = Bonferroni corrected *P*-value based on the number of genes tested per tissue | | | | |  |
| (*P*=0.05/n where n = number of genes tested) | | | | |  |

**Supplementary Table 8.** Mendelian Randomization results with depression as the exposure and type 2 diabetes as the outcome variable.

Legend: IVW=Inverse variance weighted, **Nsnps** = Number of SNPs (instrument variables) used, **OR(95%CI)** = odds ratio and lower and upper 95% confidence intervals (CI)

| **Method** | **Outcome** | **Exposure** | **N_SNPs_** | **OR (95%CI)** | ***P*** |
| --- | --- | --- | --- | --- | --- |
| IVW | Type 2 Diabetes | Depression | 95 | 1.18 (1.061-1.32) | 0.0024 |
| Weighted median | Type 2 Diabetes | Depression | 95 | 1.11 (1.00-1.23) | 0.043 |
| MR Egger | Type 2 Diabetes | Depression | 95 | 0.95 (0.50-1.81) | 0.88 |
| Simple mode | Type 2 Diabetes | Depression | 95 | 0.89 (0.60-1.32) | 0.56 |
| Weighted mode | Type 2 Diabetes | Depression | 95 | 1.29 (0.89-1.87) | 0.18 |

**Supplementary Table 9.** Mendelian Randomization results with type 2 diabetes as the exposure and depression as the outcome variable

| **Method** | **Outcome** | **Exposure** | **N_SNPs_** | **OR(95%CI)** | ***P*** |
| --- | --- | --- | --- | --- | --- |
| IVW | Depression | Type 2 Diabetes | 344 | 0.999 (0.986-1.011) | 0.84 |
| Weighted median | Depression | Type 2 Diabetes | 344 | 0.995 (0.979-1.011) | 0.52 |
| MR Egger | Depression | Type 2 Diabetes | 344 | 0.984 (0.959-1.009) | 0.204 |
| Simple mode | Depression | Type 2 Diabetes | 344 | 1.004 (0.964-1.045) | 0.85 |
| Weighted mode | Depression | Type 2 Diabetes | 344 | 0.988 (0.967-1.0102) | 0.29 |

Legend: IVW=Inverse variance weighted, **Nsnps** = Number of SNPs (instrument variables) used, **OR(95%CI)** = odds ratio and lower and upper 95% confidence intervals (CI)

**Supplementary Table 10.** The number of genome-wide significant independent signals for each studied phenotype, identified with each analytical approach, implemented in either BOLT-LMM or MTAG in UKBB

| Software tool | Type of GWAS | Type 2 diabetes | MDD | PHQ-9 |
| --- | --- | --- | --- | --- |
| BOLT-LMM | Single phenotype | 92 | 0 | 3 |
| MTAG (Type 2 diabetes + MDD) | Multi-phenotype | 68 | 0 | -* |
| MTAG (Type 2 diabetes + PHQ-9) | Multi-Phenotype | 53 | -* | 8 |

* “-“The phenotype is not analyzed in this model

| **Supplementary Table 11. Summary statistics for signals reaching genome-wide significance in GWAS of PHQ-9 and Type 2 diabetes in UKBB and their replication results in FinnGen.** | | | | | | | |  |  |  |  |  |
| --- | --- | --- | --- | --- | --- | --- | --- | --- | --- | --- | --- | --- |
| **CHR** | **BP** | **SNP** | **NEAREST**  **GENE** | **EA** | **NEA** | **EAF** | **BETA** | **SE** | ***P*** | **BETA** | **SE** | ***P*** |
| **PHQ-9 in UK BIOBANK** | | | | | | | | | | **PHQ-9 in FINNGEN** | | |
| 1 | 201,861,016 | rs2279681 | *SHISA4* | G | C | 0.34 | -0.083 | 0.014 | 2.60x-09 | NA | NA | NA |
| 3 | 50,535,195 | rs35335661 | *CACNA2D2* | A | C | 0.13 | 0.125 | 0.020 | 1.40x-10 | NA | NA | NA |
| 8 | 106,012,986 | rs35844112 | *LRP12/ZFPM2* | T | A | 0.34 | -0.076 | 0.012 | 4.50x-08 | NA | NA | NA |
| **Type 2 diabetes in UK BIOBANK** | | | | | | | | | | **Type 2 diabetes in FINNGEN** | | |
| 1 | 40,035,686 | rs17513135 | *PABPC4/PABPC4-AS1* | T | C | 0.220 | 0.0029 | 0.0005 | 6.40x-10 | 0.061 | 0.018 | 0.000975 |
| 1 | 120,461,253 | rs2793829 | *NOTCH2* | T | C | 0.113 | 0.0038 | 0.0006 | 4.80x-10 | 0.064 | 0.023 | 0.0047 |
| 1 | 146,747,153 | rs6680778 | *CHD1L/NBPF19* | C | T | 0.858 | -0.0032 | 0.0006 | 1.10x-08 | 0.075 | 0.020 | 0.000175 |
| 1 | 214,159,256 | rs340874 | *PROX1-AS1* | C | T | 0.558 | 0.0023 | 0.0004 | 2.60x-09 | 0.040 | 0.016 | 0.01124 |
| 1 | 229,672,955 | rs348330 | *ABCB10* | A | G | 0.620 | -0.0022 | 0.0004 | 4.30x-08 | -0.054 | 0.016 | 0.000888 |
| 2 | 25,513,652 | rs6739187 | *DNMT3A* | A | G | 0.431 | -0.0022 | 0.0004 | 2.60x-08 | -0.017 | 0.016 | 0.2761 |
| 2 | 27,730,940 | rs1260326 | *GCKR* | C | T | 0.612 | 0.0025 | 0.0004 | 3.60x-10 | 0.045 | 0.016 | 0.005359 |
| 2 | 43,575,984 | rs13411485 | *THADA* | G | C | 0.122 | -0.0043 | 0.0006 | 4.30x-13 | -0.040 | 0.032 | 0.2048 |
| 2 | 60,586,707 | rs243018 | *MIR4432HG* | G | C | 0.456 | 0.0025 | 0.0004 | 1.90x-10 | 0.043 | 0.016 | 0.006128 |
| 2 | 121,347,612 | rs11688682 | *GLI2* | C | G | 0.265 | -0.0026 | 0.0005 | 2.20x-08 | -0.032 | 0.018 | 0.07997 |
| 2 | 165,528,876 | rs13389219 | *GRB14/COBLL1* | T | C | 0.395 | -0.0039 | 0.0004 | 4.80x-22 | -0.069 | 0.016 | 2.19x-05 |
| 2 | 227,101,411 | rs2972144 | *IRS1* | G | A | 0.653 | 0.0036 | 0.0004 | 1.70x-18 | 0.068 | 0.016 | 2.07x-05 |
| 3 | 12,386,337 | rs4684847 | *PPARG* | T | C | 0.117 | -0.0047 | 0.0006 | 9.70x-15 | -0.102 | 0.021 | 1.03x-06 |
| 3 | 23,454,790 | rs1496653 | *MIR548AC* | G | A | 0.208 | -0.0033 | 0.0005 | 7.90x-12 | -0.064 | 0.017 | 0.000131 |
| 3 | 123,069,058 | rs11720108 | *ADCY5* | T | C | 0.243 | -0.0041 | 0.0005 | 3.10x-19 | -0.095 | 0.020 | 3.11x-06 |
| 3 | 170,663,148 | rs2422133 | *SLC2A2* | G | A | 0.398 | -0.0032 | 0.0004 | 6.50x-16 | -0.044 | 0.016 | 0.005445 |
| 3 | 185,513,296 | rs7640539 | *IGF2BP2* | A | T | 0.321 | 0.0052 | 0.0004 | 1.10x-35 | 0.079 | 0.017 | 2.59x-06 |
| 3 | 186,665,645 | rs3887925 | *ST6GAL1* | T | C | 0.542 | 0.0024 | 0.0004 | 5.80x-10 | 0.071 | 0.016 | 5.80x-06 |
| 3 | 187,740,899 | rs4686471 | *LPP-AS2* | C | T | 0.617 | 0.0026 | 0.0004 | 1.70x-10 | 0.033 | 0.016 | 0.03642 |
| 4 | 727,695 | rs73221126 | *PCGF3* | C | A | 0.040 | 0.0061 | 0.0010 | 4.80x-10 | 0.056 | 0.035 | 0.1096 |
| 4 | 1,784,403 | rs56337234 | *TACC3/FGFR3* | T | C | 0.492 | -0.0025 | 0.0004 | 2.10x-10 | -0.027 | 0.016 | 0.08878 |
| 4 | 6,302,519 | rs1801212 | *WFS1* | A | G | 0.730 | 0.0036 | 0.0004 | 2.40x-16 | 0.099 | 0.016 | 1.90x-09 |
| 4 | 157,680,199 | rs12641088 | *CTSO.PDGFC* | T | C | 0.334 | -0.0023 | 0.0004 | 4.50x-08 | -0.026 | 0.018 | 0.1465 |
| 5 | 14,753,745 | rs17250977 | *ANKH* | G | A | 0.039 | 0.0059 | 0.0010 | 4.40x-09 | 0.012 | 0.055 | 0.8308 |
| 5 | 14,768,092 | rs6885132 | *ANKH* | G | C | 0.106 | -0.0037 | 0.0006 | 7.90x-09 | -0.051 | 0.024 | 0.03569 |
| 5 | 53,276,301 | rs1664781 | *ARL15* | A | G | 0.697 | 0.0024 | 0.0004 | 2.20x-08 | -0.010 | 0.016 | 0.5337 |
| 5 | 55,806,751 | rs459193 | *C5orf67* | G | A | 0.739 | 0.0031 | 0.0004 | 4.90x-12 | 0.046 | 0.017 | 0.005953 |
| 5 | 55,861,786 | rs9686661 | *C5orf67* | T | C | 0.200 | 0.0029 | 0.0005 | 2.20x-09 | 0.038 | 0.023 | 0.09161 |
| 5 | 101,714,634 | rs114546213 | *SLCO6A1* | A | G | 0.046 | 0.0065 | 0.0009 | 3.20x-12 | 0.143 | 0.037 | 0.000123 |
| 5 | 102,331,465 | rs116782923 | *PAM* | T | A | 0.051 | 0.0066 | 0.0009 | 1.20x-13 | 0.124 | 0.033 | 0.000181 |
| 6 | 7,231,843 | rs9379084 | *RREB1* | A | G | 0.114 | -0.0059 | 0.0006 | 4.90x-21 | -0.067 | 0.025 | 0.00704 |
| 6 | 20,457,100 | rs76257365 | *E2F3* | C | A | 0.039 | 0.0061 | 0.0010 | 4.20x-09 | 0.055 | 0.031 | 0.07777 |
| 6 | 20,686,573 | rs7766070 | *CDKAL1* | A | C | 0.263 | 0.0057 | 0.0004 | 3.90x-38 | 0.131 | 0.016 | 9.66x-16 |
| 6 | 21,053,671 | rs114259421 | *CDKAL1* | C | G | 0.021 | 0.0083 | 0.0014 | 9.70x-09 | -0.009 | 0.036 | 0.7926 |
| 6 | 32,473,454 | rs111940905 | *HLA-DRA/HLA-DRB5* | G | A | 0.481 | 0.0023 | 0.0004 | 7.30x-09 | NA | NA | NA |
| 6 | 32,495,213 | rs35556553 | *HLA-DRB5* | T | C | 0.469 | 0.0022 | 0.0004 | 3.40x-08 | NA | NA | NA |
| 6 | 43,758,873 | rs6905288 | *VEGFA* | A | G | 0.573 | 0.0027 | 0.0004 | 5.60x-12 | 0.069 | 0.016 | 1.11x-05 |
| 6 | 126,868,567 | rs58321169 | *RSPO3* | T | C | 0.268 | 0.0030 | 0.0004 | 1.90x-11 | NA | NA | NA |
| 6 | 127,414,838 | rs719727 | *RSPO3* | G | A | 0.244 | -0.0027 | 0.0005 | 2.20x-09 | -0.026 | 0.019 | 0.1621 |
| 7 | 7,245,480 | rs56381638 | *C1GALT1* | G | A | 0.109 | 0.0035 | 0.0006 | 2.50x-08 | 0.043 | 0.0198, | 0.03193 |
| 7 | 15,064,309 | rs2191349 | *DGKB* | T | G | 0.551 | 0.0030 | 0.0004 | 1.30x-14 | 0.063 | 0.016 | 5.28x-05 |
| 7 | 28,196,413 | rs849135 | *JAZF1* | A | G | 0.488 | -0.0039 | 0.0004 | 4.20x-23 | -0.050 | 0.016 | 0.001278 |
| 8 | 41,523,745 | rs6989203 | *ANK1* | A | G | 0.241 | -0.0035 | 0.0005 | 1.60x-14 | -0.079 | 0.019 | 3.59x-05 |
| 8 | 95,960,767 | rs1320164 | *TP53INP1* | A | G | 0.511 | -0.0023 | 0.0004 | 3.70x-09 | -0.011 | 0.016 | 0.485 |
| 8 | 118,185,025 | rs3802177 | *SLC30A8* | A | G | 0.305 | -0.0044 | 0.0004 | 1.60x-25 | -0.052 | 0.016 | 0.00112 |
| 8 | 145,972,950 | rs2953845 | *ZNF251* | T | C | 0.547 | 0.0021 | 0.0004 | 3.60x-08 | 0.027 | 0.016 | 0.08742 |
| 9 | 4,291,928 | rs10974438 | *GLIS3* | C | A | 0.349 | 0.0024 | 0.0004 | 7.80x-09 | 0.048 | 0.016 | 0.002535 |
| 9 | 22,132,076 | rs2383208 | *CDKN2B-AS1* | G | A | 0.179 | -0.0062 | 0.0005 | 1.20x-34 | -0.102 | 0.022 | 2.10x-06 |
| 9 | 22,133,773 | rs76011118 | *CDKN2B-AS1* | A | G | 0.032 | 0.0086 | 0.0012 | 2.50x-12 | 0.132 | 0.043 | 0.002386 |
| 9 | 84,308,948 | rs2796441 | *TLEI* | A | G | 0.415 | -0.0030 | 0.0004 | 4.50x-14 | -0.020 | 0.016 | 0.2002 |
| 9 | 139,246,733 | rs77684335 | *GPSM1* | G | A | 0.718 | 0.0030 | 0.0004 | 4.10x-12 | NA | NA | NA |
| 10 | 12,307,894 | rs11257655 | *CDC123/CAMK1D* | T | C | 0.211 | 0.0034 | 0.0005 | 1.10x-12 | 0.0828 | 0.018 | 2.551x-06 |
| 10 | 71,332,301 | rs41277236 | *NEUROG3* | T | C | 0.042 | 0.0055 | 0.0010 | 3.00x-08 | 0.0289 | 0.051 | 0.5687 |
| 10 | 80,943,841 | rs703980 | *ZMIZ1* | A | G | 0.449 | -0.0029 | 0.0004 | 8.20x-14 | -0.0648 | 0.015 | 2.676x-05 |
| 10 | 94,467,287 | rs34744311 | *HHEX/IDE* | T | C | 0.383 | -0.0046 | 0.0004 | 3.80x-30 | -0.0659 | 0.016 | 3.232x-05 |
| 10 | 114,599,035 | rs17746147 | *TCF7L2* | G | C | 0.217 | 0.0030 | 0.0005 | 1.30x-10 | 0.0486 | 0.019 | 0.0104 |
| 10 | 114,753,553 | rs191455149 | *TCF7L2* | T | C | 0.020 | -0.0090 | 0.0014 | 1.70x-10 | -0.2017 | 0.047 | 2.059x-05 |
| 10 | 114,758,349 | rs7903146 | *TCF7L2* | T | C | 0.289 | 0.0142 | 0.0004 | 5.40x-241 | 0.2659 | 0.02 | 1.778x-41 |
| 10 | 114,904,410 | rs56260041 | *TCF7L2* | A | G | 0.205 | 0.0033 | 0.0005 | 8.20x-12 | 0.0683 | 0.023 | 0.003295 |
| 10 | 122,915,345 | rs72631105 | *FGFR2* | A | G | 0.186 | 0.0027 | 0.0005 | 4.10x-08 | 0.0377 | 0.018 | 0.03913 |
| 11 | 2,195,844 | rs10840496 | *ASCL2* | C | T | 0.627 | -0.0029 | 0.0004 | 9.80x-13 | -0.0693 | 0.017 | 2.513x-05 |
| 11 | 2,692,249 | rs231360 | *KCNQ1* | T | C | 0.403 | 0.0022 | 0.0004 | 2.20x-08 | 0.0339 | 0.016 | 0.02951 |
| 11 | 2,857,194 | rs2237895 | *KCNQ1* | C | A | 0.410 | 0.0040 | 0.0004 | 1.30x-24 | 0.0549 | 0.016 | 0.000407 |
| 11 | 17,408,404 | rs5213 | *KCNJ11* | T | C | 0.658 | -0.0034 | 0.0004 | 2.20x-16 | -0.0724 | 0.016 | 2.826x-06 |
| 11 | 69,456,000 | rs55911137 | *CCND1* | C | G | 0.026 | -0.0068 | 0.0012 | 4.30x-08 | -0.1988 | 0.11 | 0.07899 |
| 11 | 72,460,398 | rs77464186 | *ARAP1* | C | A | 0.153 | -0.0054 | 0.0005 | 2.70x-23 | -0.0881 | 0.018 | 1.246e-0 |
| 11 | 92,708,710 | rs10830963 | *MTNR1B* | G | C | 0.274 | 0.0032 | 0.0004 | 3.60x-13 | 0.0953 | 0.016 | 3.839x-09 |
| 12 | 4,384,669 | rs3217791 | *CCND2-AS1* | T | C | 0.072 | -0.0067 | 0.0008 | 8.10x-18 | NA | NA | NA |
| 12 | 4,384,844 | rs76895963 | *CCND2-AS1* | G | T | 0.020 | -0.0191 | 0.0015 | 8.20x-36 | -0.3757 | 0.049 | 1.089x-14 |
| 12 | 26,440,698 | rs12814794 | *SSPN.ITPR2* | A | G | 0.748 | -0.0025 | 0.0005 | 1.90x-08 | -0.0429 | 0.017 | 0.01238 |
| 12 | 66,252,294 | rs189339 | *HMGA2/HMGA2-AS1* | G | A | 0.851 | -0.0041 | 0.0006 | 1.40x-13 | -0.0299 | 0.023 | 0.187 |
| 12 | 66,363,070 | rs7968902 | *HMGA2* | G | T | 0.571 | 0.0026 | 0.0004 | 1.20x-10 | 0.0479 | 0.016 | 0.002347 |
| 12 | 121,429,194 | rs1169299 | *HNF1A* | C | T | 0.463 | 0.0022 | 0.0004 | 2.90x-08 | 0.0504 | 0.016 | 0.001155 |
| 12 | 124,510,391 | rs10773051 | *RFLNA* | T | C | 0.231 | -0.0026 | 0.0005 | 1.50x-08 | -0.0211 | 0.017 | 0.2119 |
| 13 | 61,611,996 | rs75200244 | *MIR3169* | C | T | 0.021 | 0.0074 | 0.0013 | 4.10x-08 | 0.1558 | 0.11 | 0.1644 |
| 13 | 80,717,156 | rs1359790 | *SPRY2* | A | G | 0.281 | -0.0039 | 0.0004 | 9.80x-20 | -0.0511 | 0.017 | 0.002657 |
| 14 | 38,848,419 | rs8017808 | *CLEC14A* | T | G | 0.253 | -0.0026 | 0.0005 | 5.40x-09 | -0.0163 | 0.017 | 0.347 |
| 15 | 38,852,386 | rs12912777 | *RASGRP1* | T | C | 0.122 | 0.0041 | 0.0006 | 3.30x-12 | 0.0543 | 0.026 | 0.03394 |
| 15 | 41,953,105 | rs28444909 | *MGA* | T | C | 0.335 | 0.0023 | 0.0004 | 3.90x-08 | 0.0127 | 0.016 | 0.4247 |
| 15 | 77,840,714 | rs11855188 | *LINGO1* | G | A | 0.736 | 0.0028 | 0.0004 | 1.80x-10 | 0.0468 | 0.018 | 0.008461 |
| 15 | 90,385,868 | rs7174644 | *AP3S2/C15orf38-AP3S2* | T | C | 0.716 | -0.0027 | 0.0004 | 3.10x-10 | -0.0379 | 0.018 | 0.0329 |
| 16 | 53,526,551 | rs9931702 | *AKTIP* | T | C | 0.463 | -0.0024 | 0.0004 | 5.30x-10 | 0.017 | 0.016 | 0.2977 |
| 16 | 75,249,170 | rs111852127 | *CTRB2/CTRB1* | A | T | 0.076 | -0.0062 | 0.0007 | 2.50x-17 | -0.1119 | 0.029 | 9.457x-05 |
| 17 | 36,101,156 | rs7501939 | *HNF1B* | C | T | 0.602 | -0.0036 | 0.0004 | 1.80x-19 | 0.0606 | 0.017 | 0.0003621 |
| 18 | 60,845,884 | rs12454712 | *BCL2* | C | T | 0.378 | -0.0026 | 0.0004 | 4.10x-11 | -0.0297 | 0.016 | 0.05952 |
| 19 | 7,243,537 | rs28376271 | *INSR* | G | A | 0.210 | 0.0028 | 0.0005 | 1.00x-08 | 0.0683 | 0.022 | 0.001506 |
| 19 | 19,419,071 | rs739846 | *SUGP1* | A | G | 0.078 | 0.0041 | 0.0007 | 2.10x-08 | 0.0567 | 0.032 | 0.07487 |
| 19 | 46,160,458 | rs35845603 | *GIPR* | G | A | 0.350 | 0.0032 | 0.0004 | 4.50x-15 | 0.0417 | 0.016 | 0.008995 |
| 19 | 46,191,828 | rs10403723 | *SNRPD2* | T | C | 0.298 | -0.0029 | 0.0004 | 6.30x-12 | -0.0305 | 0.017 | 0.07338 |
| 20 | 42,995,600 | rs35365632 | *HNF4A* | A | G | 0.105 | 0.0036 | 0.0006 | 2.20x-08 | 0.0437 | 0.022 | 0.05129 |
| 20 | 43,042,364 | rs1800961 | *HNF4A* | T | C | 0.030 | 0.0078 | 0.0011 | 4.70x-12 | 0.0001 | 0.038 | 0.9971 |
| 20 | 45,550,489 | rs4809604 | *EYA2* | G | T | 0.434 | 0.0022 | 0.0004 | 3.30x-08 | 0.0266 | 0.016 | 0.08616 |
| Legend: **CHR** = Chromosome, **SNP** = Single nucleotide polymorphism identifier, **BP** = Position in base pair, genome build 37, **EA** = Effect allele, **NEA** = Non-effect allele, **EAF** = Effect allele frequency**, BETA** =BOLT's effect size estimate for the effect allele, **SE** = Standard error of the effect size, **P** = *P*-value of association | | | | | | | | | | | | |

**Supplementary Table 12**. Signals reaching suggestive evidence of association (P<10^-6^) in GWAS of MDD in UKBB and their replication in FinnGen

| **CHR** | **BP** | **SNP** | **NEAREST GENE** | **EA** | **NEA** | **EAF** | **BETA** | **SE** | ***P*** | **BETA** | **SE** | ***P*** |
| --- | --- | --- | --- | --- | --- | --- | --- | --- | --- | --- | --- | --- |
| **MDD in UK BIOBANK** | | | | | | | | | | **MDD in FINNGEN** | | |
| 1 | 155,015,795 | rs200957796 | *DCST1* | A | G | 0.013 | 0.025 | 0.0050 | 7.10x-07 | -0.0036 | 0.062 | 0.95 |
| 6 | 7,744,840 | rs151190352 | *BMP6* | G | A | 0.013 | 0.026 | 0.0049 | 7.90x-08 | 0.14 | 0.15 | 0.36 |
| 6 | 7,757,448 | rs139046708 | *BMP6* | A | G | 0.013 | 0.026 | 0.0049 | 1.10x-07 | 0.15 | 0.15 | 0.31 |
| 10 | 11,797,186 | rs60806452 | *ECHDC3* | A | G | 0.024 | 0.019 | 0.0036 | 1.20x-07 | 0.017 | 0.037 | 0.66 |
| 11 | 12,482,646 | rs78220143 | *PARVA* | T | C | 0.016 | 0.023 | 0.0046 | 4.00x-07 | 0.022 | 0.059 | 0.71 |
| 11 | 12,515,476 | rs74421466 | *PARVA* | G | A | 0.016 | 0023 | 0.0046 | 4.90x-07 | 0.0203 | 0.063 | 0.75 |
| 11 | 55,195,318 | rs574032318 | *NA* | C | T | 0.016 | 0.025 | 0.0049 | 2.40x-07 | NA | NA | NA |
| 11 | 98,658,772 | rs115409450 | *CNTN5* | A | G | 0.020 | -0.020 | 0.0040 | 8.50x-07 | -0.043 | 0.068 | 0.53 |
| 11 | 113,810,970 | rs45452401 | *HTR3B* | T | C | 0.040 | 0.014 | 0.0028 | 6.40x-07 | -0.031 | 0.055 | 0.58 |
| 11 | 124,791,601 | rs139178836 | *HEPACAM* | T | C | 0.018 | 0.022 | 0.0042 | 1.90x-07 | -0.12 | 0.063 | 0.059 |
| 11 | 124,834,119 | rs151236242 | *CCDC15* | G | T | 0.020 | 0.020 | 0.0039 | 6.70x-07 | -0.12 | 0.062 | 0.048 |
| 11 | 124,863,641 | rs78571498 | *CCDC15* | C | A | 0.020 | 0.019 | 0.0039 | 8.90x-07 | -0.12 | 0.062 | 0.048 |
| 12 | 14,314,595 | rs9795974 | *GRIN2B/ATF7IP* | C | A | 0.675 | -0.006 | 0.0012 | 5.60x-07 | 0.0092 | 0.018 | 0.61 |
| 12 | 14,399,895 | rs112673636 | *GRIN2B/ATF7IP* | T | C | 0.259 | 0.006 | 0.0013 | 5.40x-07 | -0.0073 | 0.018 | 0.69 |
|  | Legend: **CHR** = Chromosome, **SNP** = Single nucleotide polymorphism identifier, **BP** = Position in base pair, genome build 37, **EA** = Effect allele, **NEA** = Non-effect allele, **EAF** = Effect allele frequency**, BETA** =BOLT's effect size estimate for the effect allele, **SE** = Standard error of the effect size, **P** = *P*-value of association | | | | | | | | | | | |

**Supplementary Table 13**. maxFDR for MTAG results in each model analyzed

| MTAG Model | Type 2 diabetes | MDD | PHQ-9 |
| --- | --- | --- | --- |
| Type 2 diabetes + MDD (UKBB) | 1.6% | 7.1% | -* |
| Type 2 diabetes + PHQ-9 (UKBB) | 0.98% | -* | 1.8% |
| Type 2 diabetes + MDD (FinnGen) | 11.6% | 25.5% | -* |

* “-“ The phenotype is not analyzed in this model

|  | **Supplementary Table 14**. Signals reaching genome-wide significance for type 2 diabetes in the MP-GWAS of PHQ-9 and type 2 diabetes. | | | | | | | | | | | | | |  | |  | |  | |  | |  |
| --- | --- | --- | --- | --- | --- | --- | --- | --- | --- | --- | --- | --- | --- | --- | --- | --- | --- | --- | --- | --- | --- | --- | --- |
|  | **CHR** | | **BP** | | **SNP** | | **NEAREST**  **GENE** | | **EA** | | **NEA** | | **EAF** | **BETA** | **SE** | | ***P*** | | **BETA** | | **SE** | | ***P*** |
|  | **PHQ-9 IN UK BIOBANK** | | | | | | | | | | | | | | | | | | **PHQ-9 IN FINNGEN** | | | | |
|  | 2 | | 227,101,411 | | rs2972144 | | *IRS1* | | G | | A | | 0.65 | 0.065 | 0.012 | | 1.80x10^-08^ | | NA | | NA | | NA |
|  | 3 | | 50,523,494 | | rs1467916 | | *CACNA2D2* | | G | | T | | 0.13 | 0.097 | 0.016 | | 2.59x10^-09^ | | NA | | NA | | NA |
|  | 3 | | 185,512,361 | | rs16860235 | | *IGF2BP2* | | A | | G | | 0.28 | 0.103 | 0.012 | | 4.18x10^-17^ | | NA | | NA | | NA |
|  | 6 | | 20,694,884 | | rs2206734 | | *CDKAL1* | | G | | C | | 0.18 | 0.103 | 0.014 | | 1.24x10^-12^ | | NA | | NA | | NA |
|  | 9 | | 22,134,253 | | rs10811662 | | *CDKN2B-AS1* | | A | | G | | 0.17 | -0.086 | 0.015 | | 4.31x10^-09^ | | NA | | NA | | NA |
|  | 10 | | 114,758,349 | | rs7903146 | | *TCF7L2* | | T | | C | | 0.29 | 0.207 | 0.012 | | 1.68x10^-64^ | | NA | | NA | | NA |
|  | 12 | | 4,384,844 | | rs76895963 | | *CCND2-AS1* | | G | | T | | 0.02 | -0.290 | 0.043 | | 1.91x10^-11^ | | NA | | NA | | NA |
|  | 13 | | 80,717,156 | | rs1359790 | | *SPRY2* | | A | | G | | 0.28 | -0.076 | 0.012 | | 5.91x10^-10^ | | NA | | NA | | NA |
|  | **T2D IN UK BIOBANK** | | | | | | | | | | | | | | | | | | **T2D IN FINNGEN** | | | | |
| 2 | | 25,512,438 | | rs12999687 | | *DNMT3A* | | G | | T | | 0.508 | | -0.002 | | 0.0004 | 7.28x10^-09^ | -0.02 | | 0.02 | | 0.341 | |
| 2 | | 165,528,876 | | rs13389219 | | *GRB14,COBLL1* | | T | | C | | 0.605 | | -0.003 | | 0.0004 | 2.52x10^-16^ | -0.07 | | 0.02 | | 5.30x10^-05^ | |
| 2 | | 227,101,411 | | rs2972144 | | *IRS1* | | G | | A | | 0.347 | | 0.003 | | 0.0004 | 4.89x10^-16^ | 0.07 | | 0.02 | | 5.18x10^-05^ | |
| 3 | | 12,386,337 | | rs4684847 | | *PPARG* | | T | | C | | 0.883 | | -0.004 | | 0.0006 | 2.14x10^-10^ | -0.101 | | 0.02 | | 2.49x10^-06^ | |
| 3 | | 23,454,790 | | rs1496653 | | *MIR548AC* | | G | | A | | 0.792 | | -0.003 | | 0.0004 | 5.06x10^-11^ | -0.06 | | 0.02 | | 2.50x10^-04^ | |
| 3 | | 123,065,778 | | rs11708067 | | *ADCY5* | | G | | A | | 0.761 | | -0.003 | | 0.0004 | 7.51x10^-12^ | -0.1 | | 0.02 | | 3.04x10^-05^ | |
| 3 | | 170,628,132 | | rs6769079 | | *SLC2A2* | | G | | A | | 0.625 | | -0.003 | | 0.0004 | 5.99x10^-12^ | NA | | NA | | NA | |
| 3 | | 185,512,361 | | rs16860235 | | *IGF2BP2* | | A | | G | | 0.716 | | 0.005 | | 0.0004 | 1.41x10^-30^ | NA | | NA | | NA | |
| 3 | | 186,665,645 | | rs3887925 | | *ST6GAL1* | | T | | C | | 0.458 | | 0.002 | | 0.0004 | 8.55x10^-10^ | 0.07 | | 0.02 | | 2.06x10^-05^ | |
| 3 | | 187,740,899 | | rs4686471 | | *LPP-AS2* | | C | | T | | 0.383 | | 0.002 | | 0.0004 | 6.81x10^-09^ | 0.03 | | 0.02 | | 0.042 | |
| 4 | | 1,784,403 | | rs56337234 | | *TACC3,FGFR3* | | T | | C | | 0.508 | | -0.002 | | 0.0004 | 3.89x10^-08^ | -0.03 | | 0.02 | | 9.00x10^-02^ | |
| 4 | | 6,281,458 | | rs11727100 | | *WFS1* | | C | | A | | 0.272 | | 0.003 | | 0.0004 | 8.96x10^-12^ | 0.09 | | 0.02 | | 3.41x10^-08^ | |
| 5 | | 55,806,751 | | rs459193 | | *C5orf67* | | G | | A | | 0.261 | | 0.002 | | 0.0004 | 2.15x10^-08^ | 0.04 | | 0.03 | | 0.011 | |
| 5 | | 101,714,634 | | rs114546213 | | *SLCO6A1* | | A | | G | | 0.954 | | 0.006 | | 0.0009 | 2.41x10^-10^ | NA | | NA | | NA | |
| 5 | | 102,301,358 | | rs74567345 | | *PAM* | | C | | T | | 0.951 | | 0.006 | | 0.0009 | 6.75x10^-11^ | 0.12 | | 0.03 | | 0.00025 | |
| 6 | | 7,231,843 | | rs9379084 | | *RREB1* | | A | | G | | 0.886 | | -0.005 | | 0.0006 | 1.84x10^-16^ | -0.07 | | 0.03 | | 0.01 | |
| 6 | | 20,686,573 | | rs7766070 | | *CDKAL1* | | A | | C | | 0.737 | | 0.005 | | 0.0004 | 7.79x10^-29^ | 0.13 | | 0.02 | | 1.34x10^-14^ | |
| 6 | | 32,495,213 | | rs35556553 | | *HLA-DRB5* | | T | | C | | 0.531 | | 0.002 | | 0.0004 | 3.17x10^-08^ | NA | | NA | | NA | |
| 6 | | 32,554,885 | | rs113520250 | | *HLA-DRB1* | | T | | C | | 0.66 | | 0.002 | | 0.0004 | 9.05x10^-09^ | NA | | NA | | NA | |
| 6 | | 43,758,873 | | rs6905288 | | *VEGFA* | | A | | G | | 0.427 | | 0.002 | | 0.0004 | 1.49x10^-08^ | 0.07 | | 0.02 | | 1.84x10^-05^ | |
| 7 | | 15,064,255 | | rs2191348 | | *DGKB* | | T | | G | | 0.448 | | 0.003 | | 0.0004 | 1.84x10^-12^ | 0.06 | | 0.02 | | 0.00014 | |
| 7 | | 28,196,413 | | rs849135 | | *JAZF1* | | A | | G | | 0.512 | | -0.003 | | 0.0004 | 4.34x10^-16^ | -0.05 | | 0.02 | | 0.0014 | |
| 7 | | 114,632,177 | | rs4730661 | | *MDFIC* | | A | | G | | 0.573 | | -0.004 | | 0.0007 | 9.77x10^-09^ | 0.01 | | 0.02 | | 0.389 | |
| 8 | | 41,528,178 | | rs3802315 | | *ANK1* | | T | | G | | 0.759 | | -0.003 | | 0.0004 | 1.77x10^-09^ | -0.08 | | 0.02 | | 5.74x10^-05^ | |
| 8 | | 95,960,767 | | rs1320164 | | *TP53INP1* | | A | | G | | 0.489 | | -0.002 | | 0.0004 | 2.29x10^-09^ | -0.01 | | 0.02 | | 0.482 | |
| 8 | | 118,185,025 | | rs3802177 | | *SLC30A8* | | A | | G | | 0.695 | | -0.004 | | 0.0004 | 4.29x10^-19^ | -0.05 | | 0.02 | | 0.0018 | |
| 9 | | 22,003,367 | | rs1063192 | | *CDKN2B-AS1* | | A | | G | | 0.428 | | 0.002 | | 0.0004 | 3.38x10^-08^ | 0.06 | | 0.02 | | 0.00059 | |
| 9 | | 22,134,253 | | rs10811662 | | *CDKN2B-AS1* | | A | | G | | 0.827 | | -0.005 | | 0.0005 | 7.88x10^-25^ | -0.11 | | 0.02 | | 1.58x10^-06^ | |
| 9 | | 84,308,948 | | rs2796441 | | *TLEI* | | A | | G | | 0.585 | | -0.003 | | 0.0004 | 7.7x10^-12^ | -0.02 | | 0.02 | | 0.214 | |
| 9 | | 139,246,739 | | rs58602428 | | *GPSM1* | | A | | G | | 0.282 | | 0.003 | | 0.0004 | 7.48x10^-12^ | NA | | NA | | NA | |
| 10 | | 12,307,894 | | rs11257655 | | *CDC123,CAMK1D* | | T | | C | | 0.789 | | 0.002 | | 0.0004 | 2.52x10^-08^ | 0.08 | | 0.02 | | 6.30x10^-06^ | |
| 10 | | 94,467,287 | | rs34744311 | | *HHEX/IDE* | | T | | C | | 0.617 | | -0.004 | | 0.0004 | 1.98x10^-21^ | -0.07 | | 0.02 | | 5.66x10^-05^ | |
| 10 | | 114,753,553 | | rs191455149 | | *TCF7L2* | | T | | C | | 0.98 | | -0.008 | | 0.0013 | 7.3x10^-09^ | -0.2 | | 0.05 | | 2.92x10^-05^ | |
| 10 | | 114,758,349 | | rs7903146 | | *TCF7L2* | | T | | C | | 0.711 | | 0.011 | | 0.0004 | 6.9x10^-178^ | 0.26 | | 0.02 | | 1.42x10^-38^ | |
| 10 | | 114,904,410 | | rs56260041 | | *TCF7L2* | | A | | G | | 0.795 | | 0.003 | | 0.0005 | 6.85x10^-10^ | 0.07 | | 0.02 | | 0.0035 | |
| 10 | | 122,915,345 | | rs72631105 | | *FGFR2* | | A | | G | | 0.814 | | 0.003 | | 0.0005 | 1.1x10^-08^ | 0.04 | | 0.02 | | 0.04 | |
| 11 | | 2,195,844 | | rs10840496 | | *ASCL2* | | C | | T | | 0.373 | | -0.002 | | 0.0004 | 3.84x10^-10^ | -0.07 | | 0.02 | | 6.30x10^-05^ | |
| 11 | | 2,857,194 | | rs2237895 | | *KCNQ1* | | C | | A | | 0.59 | | 0.003 | | 0.0004 | 1.63x10^-15^ | 0.05 | | 0.02 | | 0.0006 | |
| 11 | | 17,408,404 | | rs5213 | | *KCNJ11* | | T | | C | | 0.342 | | -0.003 | | 0.0004 | 2.41x10^-13^ | -0.07 | | 0.02 | | 8.76x10^-6^ | |
| 11 | | 72,460,398 | | rs77464186 | | *ARAP1* | | C | | A | | 0.847 | | -0.004 | | 0.0005 | 6.49x10^-16^ | -0.08 | | 0.02 | | 2.37x10^-06^ | |
| 12 | | 4,384,669 | | rs3217791 | | *CCND2-AS1* | | T | | C | | 0.928 | | -0.005 | | 0.0007 | 9.06x10^-14^ | NA | | NA | | NA | |
| 12 | | 4,384,844 | | rs76895963 | | *CCND2-AS1* | | G | | T | | 0.98 | | -0.015 | | 0.0014 | 3.08x10^-27^ | -0.37 | | 0.05 | | 9.90x10^-14^ | |
| 12 | | 66,252,294 | | rs189339 | | *LOC100129940* | | G | | A | | 0.149 | | -0.003 | | 0.0005 | 4.17x10^-11^ | -0.03 | | 0.02 | | 0.2 | |
| 13 | | 80,717,156 | | rs1359790 | | *SPRY2* | | A | | G | | 0.719 | | -0.003 | | 0.0004 | 8.54x10^-18^ | -0.05 | | 0.02 | | 0.003 | |
| 15 | | 38,852,386 | | rs12912777 | | *RASGRP1* | | T | | C | | 0.878 | | 0.003 | | 0.0006 | 3.66x10^-10^ | 0.05 | | 0.03 | | 0.04 | |
| 15 | | 77,840,714 | | rs11855188 | | *LINGO1* | | G | | A | | 0.264 | | 0.002 | | 0.0004 | 1.81x10^-09^ | 0.05 | | 0.02 | | 0.01 | |
| 16 | | 53,437,201 | | rs8056923 | | *RBL2* | | G | | A | | 0.507 | | -0.002 | | 0.0004 | 9.42x10^-09^ | -0.003 | | 0.02 | | 0.842 | |
| 16 | | 75,240,883 | | rs72802352 | | *CTRB2* | | A | | G | | 0.924 | | -0.005 | | 0.0007 | 9.9x10^-14^ | -0.12 | | 0.03 | | 9.42x10^-05^ | |
| 17 | | 36,101,156 | | rs7501939 | | *HNF1B* | | C | | T | | 0.398 | | -0.003 | | 0.0004 | 3.9x10^-15^ | 0.06 | | 0.02 | | 0.0006 | |
| 18 | | 60,845,884 | | rs12454712 | | *BCL2* | | C | | T | | 0.622 | | -0.002 | | 0.0004 | 1.15x10^-08^ | -0.03 | | 0.02 | | 0.08 | |
| 19 | | 7,243,537 | | rs28376271 | | *INSR* | | G | | A | | 0.79 | | 0.003 | | 0.0005 | 2.03x10^-08^ | 0.07 | | 0.02 | | 0.003 | |
| 19 | | 46,178,661 | | rs2238689 | | *GIPR* | | C | | T | | 0.596 | | 0.002 | | 0.0004 | 1.15x10^-10^ | NA | | NA | | NA | |
| 20 | | 43,042,364 | | rs1800961 | | *HNF4A* | | T | | C | | 0.97 | | 0.006 | | 0.0011 | 1.52x10^-08^ | -0.002 | | 0.04 | | 0.95 | |
|  | Legend: CHR = Chromosome, SNP = Single nucleotide polymorphism identifier, BP = Position in base pair, genome build 37, EA = Effect allele, NEA = Non-effect allele, EAF = Effect allele frequency, BETA = MTAG's effect size estimate for the effect allele, SE = Standard error of the effect size, P = *P*-value of association | | | | | | | | | | | | | | | | | | | | | | |

| **Supplementary Table 15.** Table showing T2D and PHQ9 significant eQTL genes after MP-GWAS and common genes between the two phenotyper per tissue | | | |
| --- | --- | --- | --- |
| **Tissue** | **T2D Genes** | **PHQ9 Genes** | **Common** |
| Adipose subcutaneous | *RP11-395N3.2, IRS1,NCR3LG1,WFS1,C2, ZDHHC6, EIF2S2P3, JAZF1, DPP7,HLA-DRA,AKTIP, ANK1, RBL2, AP3S2, CCDC92, LONRF1, , FGFR1OP, ITFG3, ATP13A1, ZNF664, EYA1, HLA-DRB6, DMGDH, HLA-QA2, OXCT2P1, HMG20A, BHMT* | *RP11-395N3.2, IRS1, LINC00674, NCR3LG1* | *RP11-395N3.2, IRS1, NCR3LG1* |
| Adrenal gland | *AKTIP, AP3S2, RBL2, SNUPN* |  |  |
| Amygdala | *AP3S2, HLA-DRB6, HSPA1B, SNAP47* | *HSPA1B, RNF123* | *HSPA1B* |
| Anterior cingulate cortex | *SRCIN1* |  |  |
| Frontal cortex | *CDKAL1, RBL2, SPPL3, AP3S2, HLA-DRB6* | *CDKAL1* | *CDKAL1* |
| Hippocampus | *AP3S2, HLA-DRB1, RBL2, OXCT2P1, TCF19* | |  |
| Hypothalamus | *EIF2S2P3, RBL2* | *HEMK1, EIF2S2P3* | *EIF2S2P3* |
| Liver | *RBL2, AP3S2, HLA-DRB6, NOTCH2, HLA-DQA2, MSH5, PDIK1L* |  |  |
| Pancrease | *JAZF1, ST6GAL1, RBL2, AP3S2, HLA-DRB1, SNUPN, PPARG, HLA-DQA2, HLA-DRB6, NOTCH2* |  |  |
| Putamen basal ganglia | *HLA-DRB6, HLA-DQA2* |  |  |
| Skeletal muscle | *JAZF1, RP11-307C19.2, ANK1, WFS1, HLA-DRA, PABPC4, MAN2C1, HLA-DRB6, BHMT, TOM1L2, MARK2P9* | *HEMK1, RP11-307C19.2, HLA-DRA* | *RP11-395N3.2, HLA-DRA* |
| Substantia nigra | *BET1L* | *BET1L* | *BET1L* |
| Whole blood | *EIF2S2P3, RBL2, ST6GAL1, HLA-DRB1, MIEF2, RPL22L1, CAMK1D, HSPA1B, SNUPN, SREBF1* | *EIF2S2P3, HLA-DRB1* | *EIF2S2P3, HLA-DRB1* |
| Legend: Blank cells indicate no significant genes for that tissue | | |  |

**Supplementary Table 16.** Table showing type 2 diabetes MetaXcan eQTL results for the significant genes in the tissues analyzed after MP-GWAS with PHQ-9

| **Gene** | **Z score** | **Effect size** | ***P* value** | **Var g** | **Pred perf r2** | **Pred perf pval** | **Pred perf qval** | **No of SNPs used** | **No of SNPs in cov** | **No of SNPs in model** |
| --- | --- | --- | --- | --- | --- | --- | --- | --- | --- | --- |
| **ADIPOSE SUBCUTANEOUS** | | | | | | | | | | |
| *RP11-395N3.2* | -7.92 | -0.0136 | 2.36e-15 | 0.021 | 0.0729 | 7.5e-07 | NA | 9 | 9 | 9 |
| *IRS1* | -7.36 | -0.00587 | 1.86e-13 | 0.0956 | 0.106 | 9.75e-10 | NA | 34 | 34 | 34 |
| *NCR3LG1* | -6.92 | -0.0128 | 4.4e-12 | 0.0178 | 0.072 | 6.57e-07 | NA | 4 | 4 | 4 |
| *ZDHHC6* | -6.81 | -0.00574 | 9.97e-12 | 0.0953 | 0.0128 | 0.0417 | NA | 76 | 77 | 77 |
| *EIF2S2P3* | -6.71 | -0.0368 | 2,00E-11 | 0.00247 | 0.013 | 0.0416 | NA | 2 | 2 | 2 |
| *JAZF1* | -6.36 | -0.00609 | 1.98e-10 | 0.0753 | 0.0683 | 1.5e-06 | NA | 48 | 48 | 48 |
| *WFS1* | 5.94 | 0.0132 | 2.82e-09 | 0.0131 | 0.0175 | 0.0149 | NA | 21 | 21 | 21 |
| *AKTIP* | 5.36 | 0.0176 | 8.29e-08 | 0.0064 | 0.0244 | 0.00508 | NA | 16 | 16 | 16 |
| *HLA-DRA* | 5.26 | 0.00552 | 1.48e-07 | 0.057 | 0.123 | 4.36e-11 | NA | 14 | 14 | 14 |
| *ANK1* | 5.19 | 0.00961 | 2.12e-07 | 0.0207 | 0.0382 | 0.000368 | NA | 16 | 16 | 16 |
| *RBL2* | 5.08 | 0.00212 | 3.68e-07 | 0.394 | 0.469 | 4.19e-50 | NA | 56 | 56 | 56 |
| *AP3S2* | 4.95 | 0.00187 | 7.44e-07 | 0.457 | 0.505 | 7.96e-58 | NA | 32 | 32 | 32 |
| *FGFR1OP* | 4.93 | 0.0134 | 8.02e-07 | 0.00843 | 0.0167 | 0.0196 | NA | 17 | 17 | 17 |
| *LONRF1* | -4.92 | -0.0133 | 8.59e-07 | 0.00816 | 0.0273 | 0.00304 | NA | 5 | 5 | 5 |
| *C2* | 4.9 | 0.00766 | 9.42e-07 | 0.0268 | 0.0203 | 0.0105 | NA | 21 | 22 | 22 |
| *CCDC92* | -4.89 | -0.0035 | 9.93e-07 | 0.135 | 0.185 | 1.07e-16 | NA | 26 | 26 | 26 |
| *ATP13A1* | 4.89 | 0.00536 | 1.01e-06 | 0.0533 | 0.119 | 1.41e-10 | NA | 11 | 11 | 11 |
| *ITFG3* | 4.83 | 0.00274 | 1.39e-06 | 0.189 | 0.177 | 6.76e-16 | NA | 52 | 52 | 52 |
| *EYA1* | 4.76 | 0.00467 | 1.93e-06 | 0.0619 | 0.142 | 9.51e-13 | NA | 11 | 11 | 11 |
| *ZNF664* | -4.71 | -0.00702 | 2.46e-06 | 0.0308 | 0.0678 | 2.06e-06 | NA | 10 | 10 | 10 |
| *HMG20A* | 4.62 | 0.0057 | 3.8e-06 | 0.045 | 0.0966 | 9.79e-09 | NA | 11 | 11 | 11 |
| *HLA-DRB6* | 4.59 | 0.0016 | 4.47e-06 | 0.533 | 0.674 | 1.38e-94 | NA | 34 | 34 | 34 |
| *DMGDH* | -4.57 | -0.0138 | 4.98e-06 | 0.00693 | 0.0172 | 0.0188 | NA | 22 | 22 | 22 |
| *BHMT* | -4.54 | -0.00271 | 5.68e-06 | 0.182 | 0.277 | 3.34e-25 | NA | 11 | 11 | 11 |
| *OXCT2P1* | 4.53 | 0.00347 | 5.84e-06 | 0.122 | 0.183 | 3.18e-16 | NA | 16 | 16 | 16 |
| **ADRENAL GLAND** | | | | | | | | | | |
| *AKTIP* | 5.29 | 0.00461 | 1.22e-07 | 0.0902 | 0.137 | 7.88e-06 | NA | 30 | 30 | 30 |
| *AP3S2* | 4.89 | 0.00193 | 9.88e-07 | 0.36 | 0.505 | 2.09e-25 | NA | 23 | 23 | 23 |
| *RBL2* | 4.57 | 0.00199 | 4.82e-06 | 0.368 | 0.41 | 2.2e-18 | NA | 71 | 71 | 71 |
| *SNUPN* | -4.47 | -0.0051 | 7.85e-06 | 0.0422 | 0.131 | 4.77e-06 | NA | 5 | 5 | 5 |
| **AMYGDALA** | | | | | | | | | | |
| *AKTIP* | 5.29 | 0.00461 | 1.22e-07 | 0.0902 | 0.137 | 7.88e-06 | NA | 30 | 30 | 30 |
| *AP3S2* | 4.89 | 0.00193 | 9.88e-07 | 0.36 | 0.505 | 2.09e-25 | NA | 23 | 23 | 23 |
| *RBL2* | 4.57 | 0.00199 | 4.82e-06 | 0.368 | 0.41 | 2.2e-18 | NA | 71 | 71 | 71 |
| *SNUPN* | -4.47 | -0.0051 | 7.85e-06 | 0.0422 | 0.131 | 4.77e-06 | NA | 5 | 5 | 5 |
| **ANTERIOR CINGULATE CORTEX** | | | | | | | | | | |
| *SRCIN1* | 4.72 | 0.00809 | 2.34e-06 | 0.0231 | 0.0528 | 0.0271 | NA | 5 | 5 | 5 |
| *MYPOP* | -4.33 | -0.00665 | 1.48e-05 | 0.0323 | 0.0663 | 0.01 | NA | 16 | 16 | 16 |
| **FRONTAL CORTEX** | | | | | | | | | | |
| *CDKAL1* | 8.08 | 0.00717 | 6.47e-16 | 0.0901 | 0.0571 | 0.0162 | NA | 45 | 45 | 45 |
| *RBL2* | 5.29 | 0.00231 | 1.25e-07 | 0.378 | 0.429 | 9.34e-14 | NA | 44 | 44 | 44 |
| *SPPL3* | -5.12 | -0.00277 | 3.07e-07 | 0.219 | 0.138 | 0.000192 | NA | 78 | 78 | 78 |
| *AP3S2* | 4.93 | 0.00479 | 8.21e-07 | 0.06 | 0.0691 | 0.00997 | NA | 13 | 13 | 13 |
| **HIPPOCAMPUS** | | | | | | | | | | |
| *AP3S2* | 4.88 | 0.0035 | 1.05e-06 | 0.114 | 0.138 | 7.81e-05 | NA | 17 | 17 | 17 |
| *HLA-DRB1* | -4.75 | -0.00607 | 2.01e-06 | 0.058 | 0.121 | 0.000167 | NA | 9 | 9 | 9 |
| *RBL2* | 4.61 | 0.00287 | 4.01e-06 | 0.181 | 0.25 | 3.17e-07 | NA | 40 | 40 | 40 |
| *TCF19* | 4.36 | 0.00494 | 1.33e-05 | 0.0557 | 0.0581 | 0.0201 | NA | 38 | 38 | 38 |
| *OXCT2P1* | 4.33 | 0.0032 | 1.48e-05 | 0.145 | 0.117 | 0.000407 | NA | 20 | 21 | 21 |
| **HYPOTHALAMUS** | | | | | | | | | | |
| *EIF2S2P3* | -5.95 | -0.00224 | 2.62e-09 | 0.488 | 0.17 | 5.95e-05 | NA | 112 | 112 | 112 |
| *RBL2* | 4.75 | 0.00254 | 1.99e-06 | 0.232 | 0.387 | 1.1e-11 | NA | 20 | 20 | 20 |
| **LIVER** | | | | | | | | | | |
| *RBL2* | 5.3 | 0.00516 | 1.15e-07 | 0.0767 | 0.121 | 5.02e-05 | NA | 23 | 23 | 23 |
| *AP3S2* | 4.95 | 0.00242 | 7.25e-07 | 0.283 | 0.42 | 1.26e-18 | NA | 18 | 18 | 18 |
| *HLA-DRB6* | 4.8 | 0.00174 | 1.56e-06 | 0.457 | 0.422 | 1.49e-17 | NA | 92 | 92 | 92 |
| *NOTCH2* | -4.65 | -0.00985 | 3.31e-06 | 0.0165 | 0.0332 | 0.0384 | NA | 22 | 22 | 22 |
| *MSH5* | 4.53 | 0.00282 | 5.97e-06 | 0.164 | 0.069 | 0.00263 | NA | 57 | 58 | 58 |
| *PDIK1L* | 4.49 | 0.00842 | 6.97e-06 | 0.0181 | 0.0863 | 0.000731 | NA | 15 | 15 | 15 |
| *HLA-DQA2* | 4.48 | 0.00179 | 7.47e-06 | 0.428 | 0.474 | 3.86e-21 | NA | 67 | 67 | 67 |
| **PANCREAS** | | | | | | | | | | |
| *JAZF1* | -6.75 | -0.00485 | 1.51e-11 | 0.121 | 0.186 | 1.06e-09 | NA | 46 | 46 | 46 |
| *RBL2* | 5.56 | 0.00242 | 2.76e-08 | 0.36 | 0.539 | 4.52e-35 | NA | 23 | 23 | 23 |
| *ST6GAL1* | 5.39 | 0.00428 | 6.99e-08 | 0.102 | 0.181 | 3.67e-09 | NA | 17 | 17 | 17 |
| *AP3S2* | 4.98 | 0.00214 | 6.43e-07 | 0.341 | 0.491 | 4.09e-29 | NA | 17 | 17 | 17 |
| *SNUPN* | -4.89 | -0.00602 | 1.01e-06 | 0.0432 | 0.0725 | 0.000375 | NA | 9 | 9 | 9 |
| *HLA-DRB1* | -4.85 | -0.00277 | 1.24e-06 | 0.193 | 0.229 | 7.72e-12 | NA | 41 | 41 | 41 |
| *PPARG* | -4.65 | -0.00573 | 3.25e-06 | 0.0481 | 0.0624 | 0.000631 | NA | 32 | 32 | 32 |
| *NOTCH2* | -4.57 | -0.00308 | 4.77e-06 | 0.166 | 0.152 | 1.02e-07 | NA | 77 | 77 | 77 |
| *HLA-DQA2* | 4.56 | 0.00208 | 5.21e-06 | 0.309 | 0.425 | 3.15e-24 | NA | 23 | 23 | 23 |
| *HLA-DRB6* | 4.45 | 0.00175 | 8.45e-06 | 0.411 | 0.516 | 3.72e-32 | NA | 47 | 49 | 49 |
| **PUTAMENS BASAL GANGLIA** | | | | | | | | | | |
| *HLA-DQA2* | 4.34 | 0.00274 | 1.4e-05 | 0.185 | 0.364 | 1.79e-11 | NA | 28 | 28 | 28 |
| *HLA-DRB6* | 4.33 | 0.00172 | 1.47e-05 | 0.429 | 0.599 | 1.25e-21 | NA | 34 | 34 | 34 |
| **SKELETAL MUSCLE** | | | | | | | | | | |
| *JAZF1* | -8.17 | -0.0101 | 2.96e-16 | 0.0415 | 0.0922 | 2.21e-10 | NA | 13 | 13 | 13 |
| *RP11-307C19.2* | 6.28 | 0.0149 | 3.46e-10 | 0.0112 | 0.0199 | 0.0041 | NA | 9 | 9 | 9 |
| *ANK1* | 6.13 | 0.0083 | 8.72e-10 | 0.039 | 0.0784 | 6.46e-09 | NA | 12 | 12 | 12 |
| *WFS1* | 5.85 | 0.00653 | 5,00E-09 | 0.0505 | 0.0897 | 4.07e-10 | NA | 9 | 9 | 9 |
| *HLA-DRA* | 5.16 | 0.0065 | 2.47e-07 | 0.0419 | 0.069 | 5.55e-08 | NA | 27 | 27 | 27 |
| *PABPC4* | -4.91 | -0.00664 | 8.98e-07 | 0.0384 | 0.0754 | 1.14e-08 | NA | 14 | 14 | 14 |
| *MAN2C1* | -4.82 | -0.00618 | 1.42e-06 | 0.0362 | 0.0965 | 8.16e-11 | NA | 7 | 7 | 7 |
| *HLA-DRB6* | 4.74 | 0.00173 | 2.15e-06 | 0.519 | 0.653 | 1.81e-113 | NA | 48 | 48 | 48 |
| *BHMT* | -4.72 | -0.00642 | 2.37e-06 | 0.0334 | 0.0693 | 4.47e-08 | NA | 17 | 17 | 17 |
| *TOM1L2* | -4.68 | -0.00247 | 2.87e-06 | 0.223 | 0.232 | 1.92e-26 | NA | 48 | 49 | 49 |
| *MARK2P9* | 4.67 | 0.00493 | 3.02e-06 | 0.055 | 0.0404 | 3.48e-05 | NA | 39 | 39 | 39 |
| **SUBSTANTIA NIGRA** | | | | | | | | | | |
| *BET1L* | -4.4 | -0.00547 | 1.11e-05 | 0.0472 | 0.074 | 0.0328 | NA | 35 | 35 | 35 |
| **WHOLE BLOOD** | | | | | | | | | | |
| *EIF2S2P3* | -8.84 | -0.0127 | 9.22e-19 | 0.032 | 0.0734 | 1.1e-06 | NA | 18 | 18 | 18 |
| *RBL2* | 5.67 | 0.00513 | 1.41e-08 | 0.0801 | 0.157 | 9.78e-14 | NA | 20 | 20 | 20 |
| *ST6GAL1* | 5.5 | 0.0115 | 3.78e-08 | 0.0159 | 0.0473 | 0.000105 | NA | 15 | 15 | 15 |
| *HLA-DRB1* | -5.34 | -0.00453 | 9.35e-08 | 0.106 | 0.159 | 7.77e-14 | NA | 21 | 21 | 21 |
| *MIEF2* | 5.01 | 0.00836 | 5.49e-07 | 0.0242 | 0.0272 | 0.00379 | NA | 21 | 22 | 22 |
| *RPL22L1* | -4.8 | -0.00343 | 1.59e-06 | 0.127 | 0.249 | 4.21e-22 | NA | 5 | 5 | 5 |
| *HSPA1B* | -4.68 | -0.0119 | 2.93e-06 | 0.0101 | 0.0193 | 0.0149 | NA | 8 | 8 | 8 |
| *CAMK1D* | 4.66 | 0.00362 | 3.12e-06 | 0.107 | 0.157 | 1.32e-13 | NA | 33 | 36 | 36 |
| *SNUPN* | -4.64 | -0.00537 | 3.56e-06 | 0.0471 | 0.0711 | 1.9e-06 | NA | 22 | 22 | 22 |
| *SREBF1* | -4.51 | -0.00474 | 6.63e-06 | 0.0593 | 0.133 | 1.63e-11 | NA | 15 | 15 | 15 |

Legend: **Z score** = MetaXcan's association result for the gene, **Effect size** = MetaXcan's association effect size for the gene, ***P* value** = *P* value of the effect size aforementioned, **Var g** = Variance of the gene expression, calculated as W' * G * W (where W is the vector of SNP weights in a gene's model, W' is its transpose, and G is the covariance matrix, **Pred perf r2** = r2 of tissue model's correlation to gene's measured transcriptome (prediction performance), **Pred perf pval** = P value of tissue model's correlation to gene's measured transcriptome (prediction performance), **Pred perf qval** = q value of tissue model's correlation to gene's measured transcriptome (prediction performance), **No of SNPs used** = number of SNPs from GWAS that got used in MetaXcan analysis, **No of SNPs in cov** = Number of SNPs in the covariance matrix, No of SNPs in model = number of SNPs in the model.

| **Gene** | **Z score** | **Effect size** | ***P* value** | **Var g** | **Pred perf r2** | **Pred perf pval** | **Pred perf qval** | **No of SNPs used** | **No of SNPs in cov** | **No of SNPs in model** |
| --- | --- | --- | --- | --- | --- | --- | --- | --- | --- | --- |
| **ADIPOSE SUBCUTANEOUS** | | | | | | | | | | |
| *RP11-395N3.2* | 5.52 | 0.29 | 3.48e-08 | 0.021 | 0.0729 | 7.5e-07 | NA | 9 | 9 | 9 |
| *IRS1* | 5.07 | 0.123 | 4.04e-07 | 0.0956 | 0.106 | 9.75e-10 | NA | 34 | 34 | 34 |
| *LINC00674* | -4.92 | -0.151 | 8.46e-07 | 0.0653 | 0.18 | 8.86e-16 | NA | 15 | 15 | 15 |
| *NCR3LG1* | 4.59 | 0.259 | 4.34e-06 | 0.0178 | 0.072 | 6.57e-07 | NA | 4 | 4 | 4 |
| *FGFR1OP* | -4.56 | -0.379 | 5.24e-06 | 0.00843 | 0.0167 | 0.0196 | NA | 17 | 17 | 17 |
| **ADRENAL GLAND** | | | | | | | | | | |
| *PSMD13* | 4.44 | 0.3 | 8.95e-06 | 0.0121 | 0.104 | 9.73e-05 | NA | 15 | 15 | 15 |
| **AMYGDALA** | | | | | | | | | | |
| *HSPA1B* | -4.59 | -0.362 | 4.5e-06 | 0.00996 | 0.0633 | 0.0317 | NA | 6 | 6 | 6 |
| *PSMD13* | 4.35 | 0.138 | 1.35e-05 | 0.0578 | 0.17 | 0.000229 | NA | 21 | 21 | 21 |
| *RNF123* | -4.26 | -0.0877 | 2.02e-05 | 0.145 | 0.128 | 0.000536 | NA | 53 | 53 | 53 |
| **FRONTAL CORTEX** | | | | | | | | | | |
| *CDKAL1* | -5.67 | -0.153 | 1.42e-08 | 0.0901 | 0.0571 | 0.0162 | NA | 45 | 45 | 45 |
| *PSMD13* | 4.4 | 0.0635 | 1.08e-05 | 0.366 | 0.395 | 1.95e-13 | NA | 44 | 44 | 44 |
| **HYPOTHALAMUS** | | | | | | | | | | |
| *HEMK1* | 5.75 | 0.327 | 9.18e-09 | 0.0304 | 0.128 | 0.000562 | NA | 9 | 9 | 9 |
| **SKELETAL MUSCLE** | | | | | | | | | | |
| *RP11-307C19.2* | -4.79 | -0.348 | 1.64e-06 | 0.0112 | 0.0199 | 0.0041 | NA | 9 | 9 | 9 |
| *HEMK1* | 4.77 | 0.134 | 1.8e-06 | 0.0872 | 0.13 | 1.43e-14 | NA | 18 | 18 | 18 |
| *HLA-DRA* | -4.6 | -0.177 | 4.22e-06 | 0.0419 | 0.069 | 5.55e-08 | NA | 27 | 27 | 27 |
| **SUBSTANTIA NIGRA** | | | | | | | | | | |
| *BET1L* | 4.97 | 0.191 | 6.54e-07 | 0.0472 | 0.074 | 0.0328 | NA | 35 | 35 | 35 |
| **WHOLE BLOOD** | | | | | | | | | | |
| *EIF2S2P3* | -5 | -0.22 | 5.87e-07 | 0.032 | 0.0734 | 1.1e-06 | NA | 18 | 18 | 18 |
| *ST6GAL1* | 4.51 | 0.287 | 6.34e-06 | 0.0159 | 0.0473 | 0.000105 | NA | 15 | 15 | 15 |
| *HLA-DRB1* | -4.49 | -0.115 | 7.03e-06 | 0.106 | 0.159 | 7.77e-14 | NA | 21 | 21 | 21 |

**Supplementary Table 17.** Table showing PHQ9 MetaXcan eQTL results for the significant genes in the tissues analyzed after MP-GWAS with type 2 diabetes

Legend: **Z score** = MetaXcan's association result for the gene, **Effect size** = MetaXcan's association effect size for the gene, ***P* value** = P value of the effect size aforementioned, **Var g** = Variance of the gene expression, calculated as W' * G * W (where W is the vector of SNP weights in a gene's model, W' is its transpose, and G is the covariance matrix, **Pred perf r2** = r2 of tissue model's correlation to gene's measured transcriptome (prediction performance), **Pred perf pval** = P value of tissue model's correlation to gene's measured transcriptome (prediction performance), **Pred perf qval** = q value of tissue model's correlation to gene's measured transcriptome (prediction performance), **No of SNPs used** = number of SNPs from GWAS that got used in MetaXcan analysis, **No of SNPs in cov** = Number of SNPs in the covariance matrix, No of SNPs in model = number of SNPs in the model.

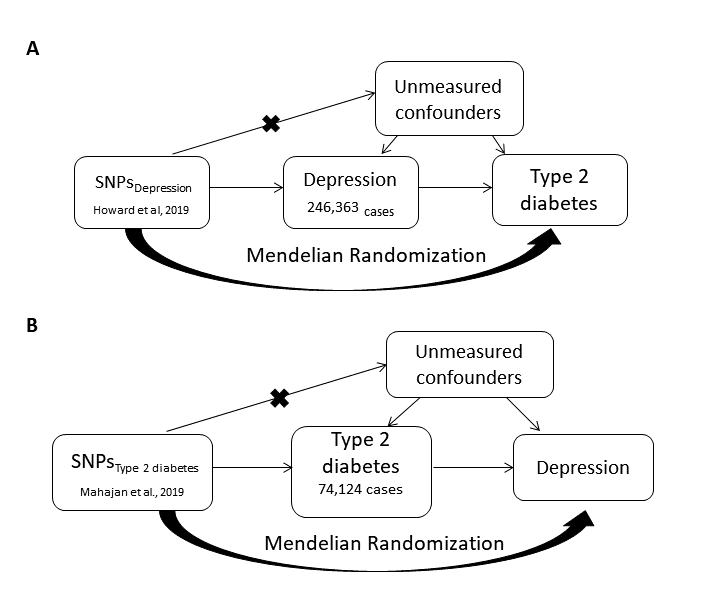

**Supplementary Figure 1**. Mendelian Randomization analysis to explore causality between depression and type 2 diabetes. (A) IV estimator is calculated as the beta coefficient from the association of GRS_depression_ with type 2 diabetes divided by the beta coefficient from the association of GRS_Type 2 diabetes_ with depression. (B) The relationship of type 2 diabetes with depression.
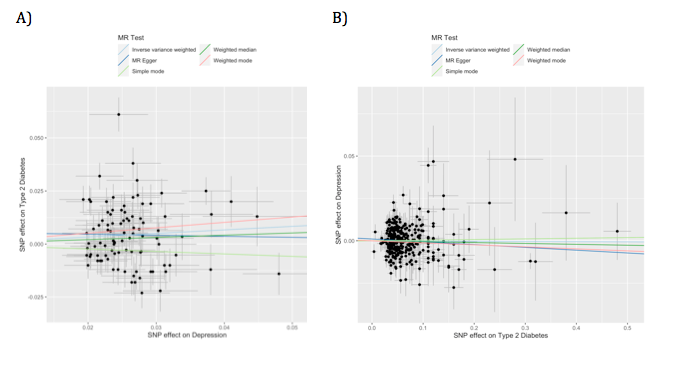

**Supplementary Figure 2.** A) Scatter plot for MR analyses of the causal effect of Depression on Type 2 Diabetes. B) Scatter plot for MR analyses of the causal effect of Type 2 Diabetes on Depression. Analyses were conducted using IVW, MR-Egger, simple mode, weighted median and weighted mode methods. The slope of each line corresponding to the estimated MR effect per method.

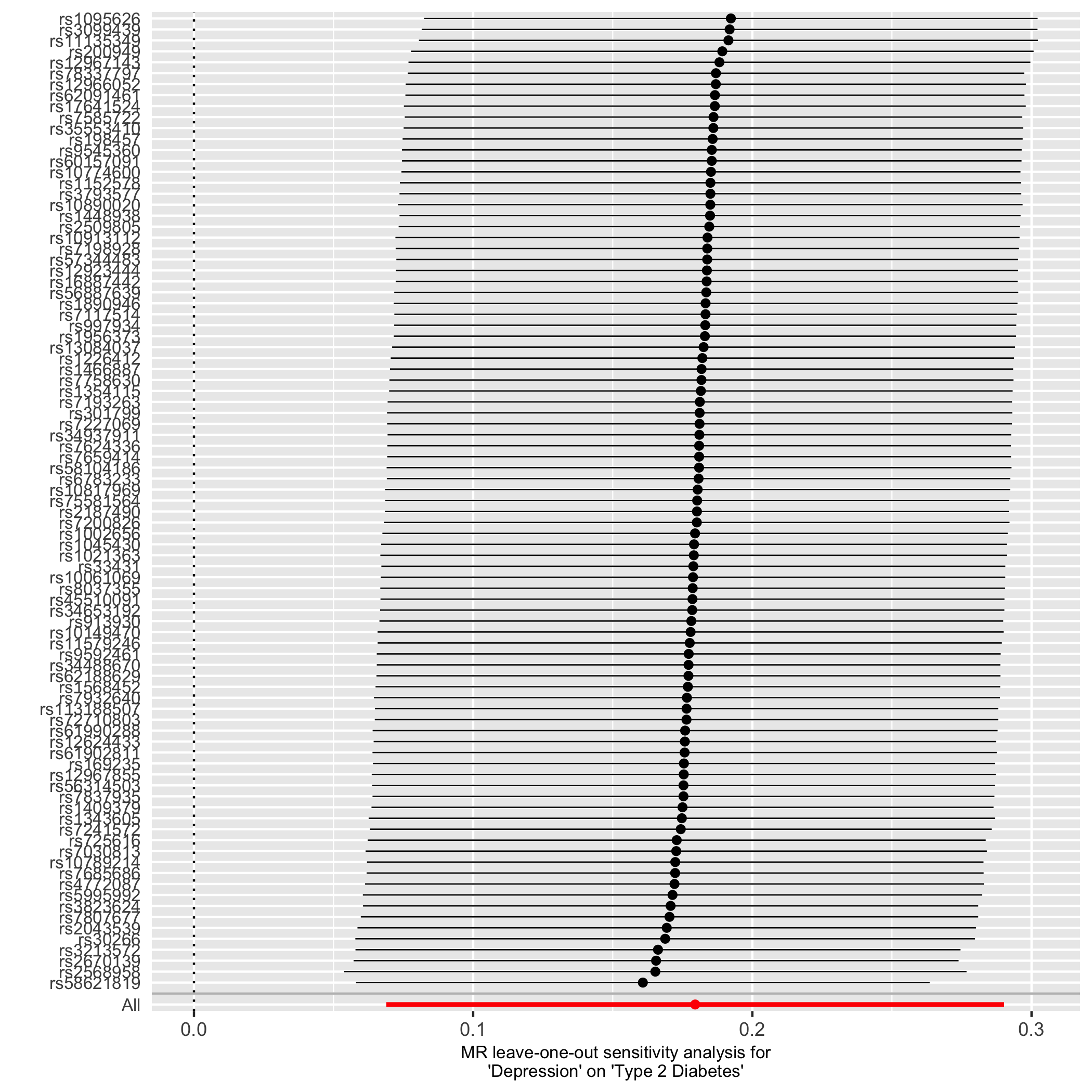

**Supplementary Figure 3.** Leave-one-out analysis: each row represents a MR analysis of Depression on Type 2 Diabetes using all instruments expect for the SNP listed on the y-axis. The point represents the odds ratio with that SNP removed and the line represents 95% confidence interval.

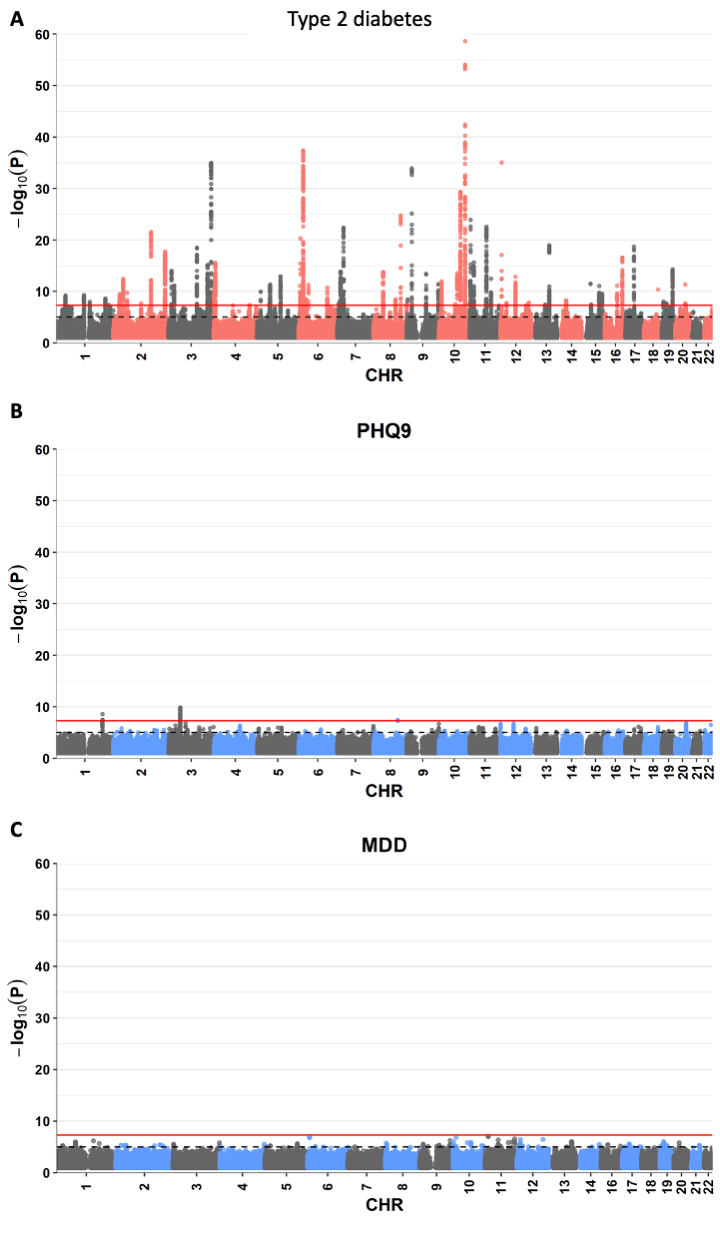

**Supplementary Figure 4**. Manhattan plots for (A) Type 2 diabetes, (B) PHQ-9 and (C) MDD SP-GWAS in the UK Biobank**.** The red horizontal line shows genome-wide significance threshold (*P*<5×10^-8^). Grey dashed horizontal lines show suggestive genome-wide significance threshold (*P*<1×10^-5^)

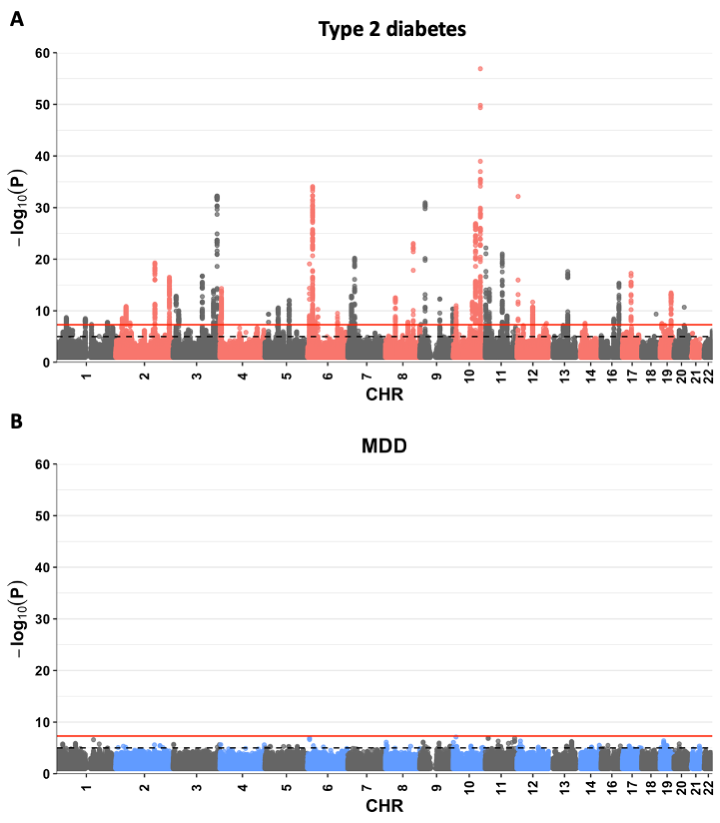
**Supplementary Figure 5**. Manhattan plots for (A) type 2 diabetes and (B) MDD after MP-GWAS in MTAG. The red horizontal line shows genome-wide significance threshold (*P*<5×10^-8^). Grey dashed horizontal lines show suggestive genome-wide significance threshold (*P*<1×10^-5^)

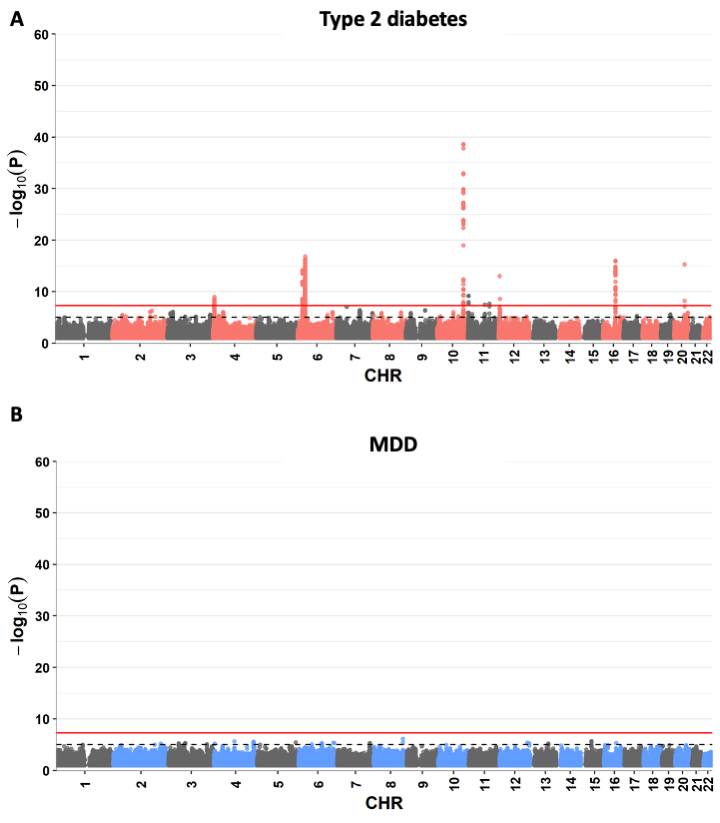
**Supplementary Figure 6**. Manhattan plots for A) type 2 diabetes and B) MDD after MP-GWAS in FinnGen dataset. Red horizontal line shows genome-wide significance threshold (*P*<5×10-8). Grey dashed horizontal lines show suggestive genome-wide significance threshold (*P*<1×10-5)

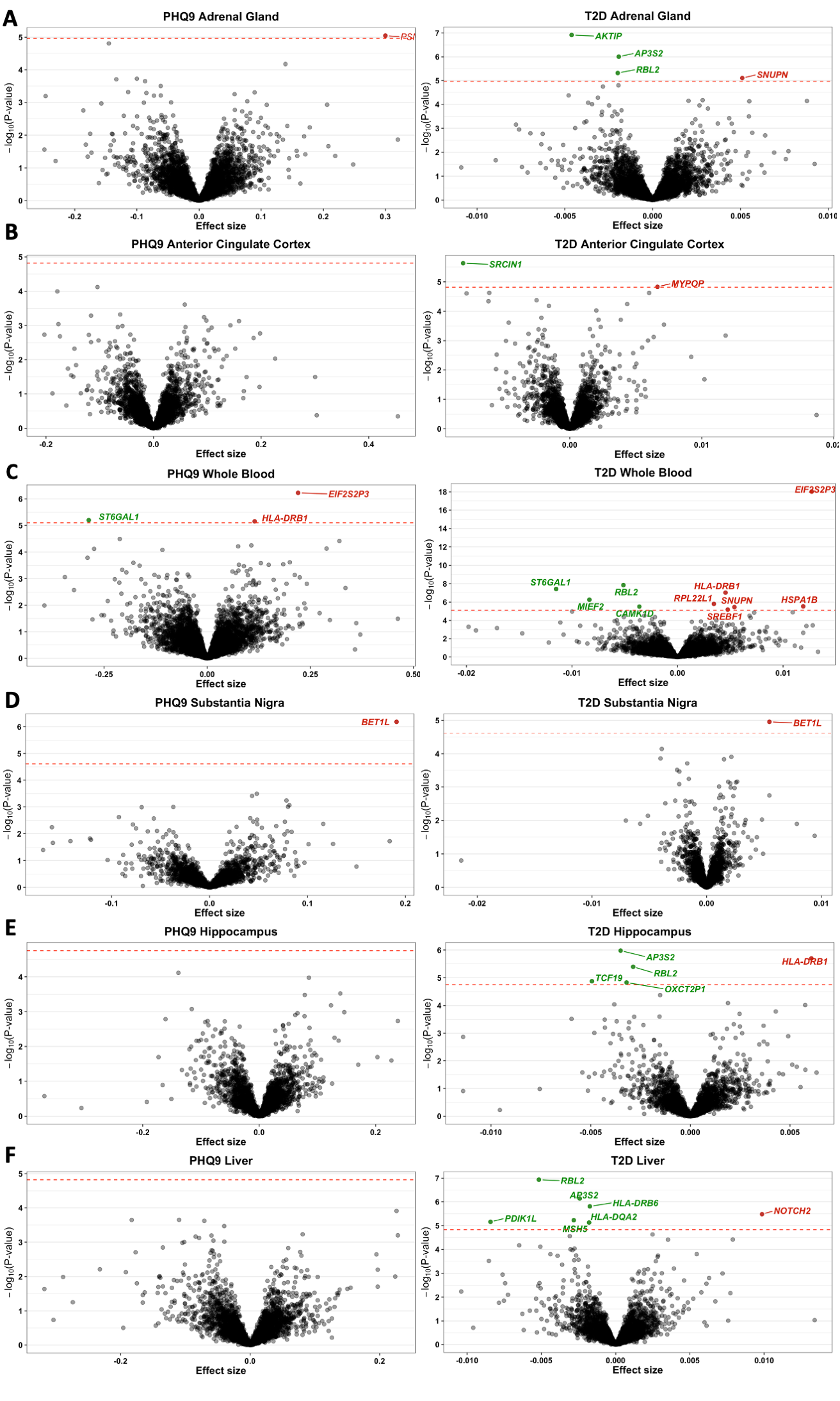

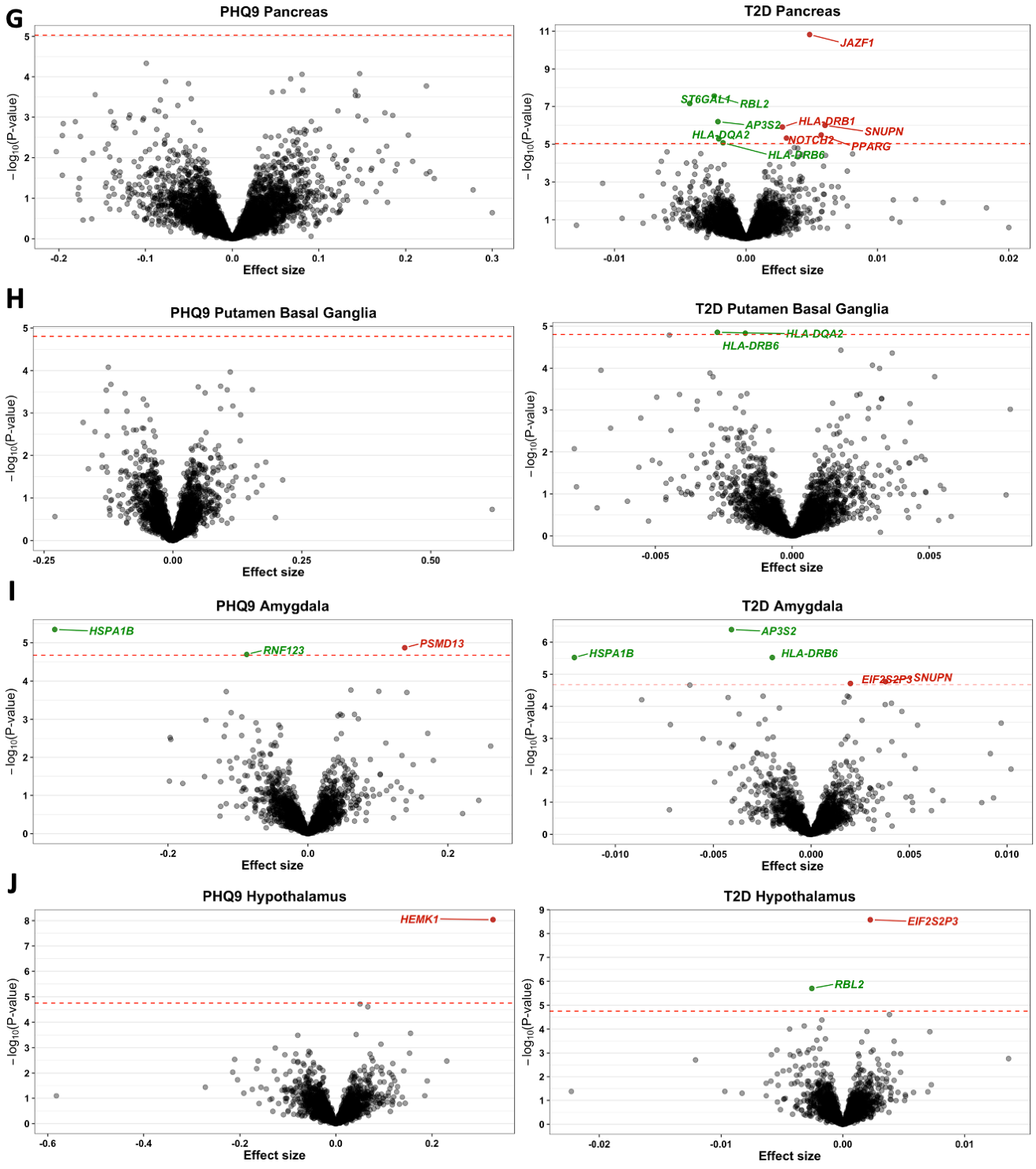

**Supplementary Figure 7.** Volcano plots of association in tissues implicated in both type 2 diabetes and PHQ-9. Each point on the plot represents an association result for one gene, where the effect size for the association of predicted gene expression and the phenotype of interest is on the x-axis and the –log10(p-value) of the association is shown on the y-axis. The red dashed line represents a Bonferroni corrected threshold for statistical significance for each tissue analyzed. Statistically significant genes for which predicted gene expression is increased are shown in red, and genes for which predicted gene expression is decreased are shown as green. Statistically insignificant genes are shown in black. A = adrenal gland, B = anterior cingulate cortex, C = whole blood, D = substantia nigra, E = hippocampus, F = liver, G = pancreas, H = putamen basal ganglia, I = amygdala, J = Hypothalamus
